# Twenty-five years of phage therapy funding: rapid growth, narrow base

**DOI:** 10.64898/2026.11.06.26367455

**Authors:** Maxime Descartes Mbogning Fonkou, Nathan Yanwou, Roland Bamou, Idriss Tsayem Ngueguim, Abdourahamane Yacouba

## Abstract

**Background:** Bacteriophage therapy is moving from compassionate use toward regulated clinical use, as antimicrobial resistance (AMR) grows. Who pays for phage therapy research, and where the money goes, has not been measured globally.

**Methods:** We searched Dimensions on 13 February 2024 and screened 1,521 grant records. We analysed the 802 grants with a recorded funding amount that started in 1999–2023 (USD 318.6 million). We estimated log-linear growth, measured concentration with the Herfindahl–Hirschman index (HHI) and Gini coefficient, and counted shared grants fractionally across recipient institutions. Topics came from latent Dirichlet allocation, and team gender from first names. Sensitivity analyses added the 272 relevant grants without an amount, merged yearly records into projects, and varied key definitions.

**Results:** Grants grew by 12.2% a year and recorded funding by 16.6% a year (14.1% after inflation), yet the field received only about USD 13 million a year. US funders provided 47.7% of funding and one institute, NIAID, 21.0%, rising to 23.3% in 2016–2023. The top 10% of recipient institutions received 53.4%. Low- and lower-middle-income countries received 0.33% of recorded funding, and only 7 grants linked Global North and Global South recipients. Antibiotic-resistance research rose from 3% to about a quarter of funding. Female-only teams received grants 19% smaller than male-only teams after adjustment (95% CI −32 to −2), and 84% of researchers held a single grant. Counting grants rather than dollars, the Global South received 17.5%, because several of its funders report no amounts. The headline results held in 37 of 40 sensitivity checks.

**Conclusions:** Phage therapy research funding is growing fast from a small base, but it rests on a narrow set of funders and countries and rarely reaches the countries most affected by AMR. Broader funding, and fuller reporting of national investments, will matter as phage therapy enters regulated clinical use.

## 1. Introduction

Antimicrobial resistance is one of the leading causes of death worldwide. In 2019, bacterial AMR was associated with an estimated 4.95 million deaths, and the burden was highest in sub-Saharan Africa and South Asia [1]. As new antibiotics reach patients slowly, interest has returned to bacteriophage therapy: the use of viruses that infect and kill specific bacteria [2,3]. Phages were used clinically before the antibiotic era and never stopped being used in Georgia, Poland and Russia. They are now being tested again in compassionate-use cases and clinical trials in Europe, North America and Australia [3,4].

The field is moving from individual cases toward regulated products. Belgium allows phages to be prepared as magistral medicines [5], and in October 2025 the European Medicines Agency published its first draft guideline on the quality of phage therapy medicinal products [6]. In 2026, the US National Institute of Allergy and Infectious Diseases (NIAID) funded three Centers for Accelerating Phage Therapy, with USD 6 million in their first year [7]. As phage therapy becomes a product rather than a last resort, who pays for the underlying research shapes which infections, which patients and which countries it will serve.

Yet no study has measured research funding for phage therapy at the global, grant level. Funding analyses exist for AMR [8,9] and for infectious disease research in general [10], but they do not separate phage therapy. We previously mapped the phage therapy patent landscape, which measured what comes out of the field as intellectual property [11]. This study measures what goes into it. Together, the two studies follow the field from money to inventions.

We ask four questions about 25 years of grants (1999–2023). How much money has gone into phage therapy research, and how fast has it grown? Who provides it, and how concentrated is it? Where does it go, by country and income group? And what, and who, gets funded, by research topic and by the gender of research teams? Because funding databases record some grants without an amount, we also test how far each answer depends on that gap.

## 2. Methods

### 2.1 Data and screening

We searched the Dimensions grants database [12] on 13 February 2024. The query combined phage terms in titles (bacteriophage, phage, phagotherapy and related forms) with therapy terms in titles or abstracts (therapy, treatment, antibacterial, antimicrobial, cocktail, prophylaxis and related forms); the full query is in the Supplementary Methods. The search returned 1,521 records.

Titles and abstracts were screened for therapeutic use of phages, which excluded 391 records and left 1,130 relevant grants (Figure 6). Of these, 312 had no funding amount in Dimensions, and 16 of the remaining 818 started outside 1999–2023. The analytic sample is the 802 grants with a recorded amount that started in 1999–2023. Dollar-based results therefore describe recorded funding. The 272 relevant grants without an amount that started in 1999–2023 were kept for sensitivity analyses (Section 2.4).

Amounts are in US dollars as converted by Dimensions. Each grant is dated by its start year. Because Dimensions indexes grants with a lag, 2023 is incomplete; it is shown in figures but excluded from trend estimates.

### 2.2 Classifications

Funders and recipient institutions were assigned a country from Dimensions’ standardised organisation records. Each country was given a World Bank income group (fiscal year 2024) and a Global North or Global South label from one reference table, used for both funders and recipients [13,14]. The table is published with the code. It documents the cases where sources disagree: Russia, Georgia and Ukraine are labelled Global North, Singapore Global South, and Taiwan, which the World Bank does not list separately, high income. The European Commission and the European Research Council are coded as funder country “European Union”.

A grant with several recipient institutions gives each an equal share of its amount and of the grant (fractional counting), so country and income-group totals add up to the whole. A grant is called domestic when every recipient institution is in the funder’s country.

### 2.3 Analyses

#### Growth

Annual growth was estimated with a log-linear regression of yearly grant counts and USD totals on year, over 1999–2022, and reported with 95% confidence intervals. Growth is also reported in constant 2023 dollars, deflated by the US consumer price index (CPI-U) by start year.

#### Concentration

We computed the HHI (the sum of squared percentage shares, 0–10,000) for funders, funder countries and recipient countries. Above 2,500 is usually called highly concentrated. For recipient institutions we report the Gini coefficient and the share received by the top 10%. US and NIAID shares are reported for 1999–2008, 2009–2015 and 2016–2023.

#### Topics

Titles, abstracts and keywords (English translations) were lemmatised with spaCy (nouns, adjectives and verbs). Words on the WordCloud and NLTK stopword lists were removed. A term-count matrix kept terms in at least 50 grants and at most 75% of them. We fitted latent Dirichlet allocation (LDA) models [15] with 2 to 12 topics and chose k = 5, where c_v coherence [16] was highest. Each grant was assigned its most probable topic. Four topics were interpretable and the fifth held a single grant, which we report as unclassified. Refitting with five other random seeds kept 51–68% of grants in the same topic, so topic results are reported as aggregate trends.

#### Gender

The gender of named researchers was inferred from first names with Gender API, with manual checks of ambiguous names [17]. Grants were placed in four exclusive groups: male only, female only, mixed, and gender unknown (no named researcher, or none whose gender could be inferred). Grant sizes were compared with Mann–Whitney tests. The main model is an ordinary least squares regression of log(amount) on team group, log team size, log grant length, start year, funder (funders with fewer than 10 grants pooled) and topic, with HC1 robust standard errors.

#### Engagement

Researchers, institutions and funders were labelled by their activity across years, as in our patent study [11]: newcomer, continuant, terminator or transient. Because these labels depend on whether a grant counts in its start year only or in every year it runs, our main measure of retention is the share of researchers who held a single grant.

### 2.4 Sensitivity analyses

We tested whether each headline result depends on the analytic sample or on a definition. First, the 272 relevant grants without an amount were classified by award type (research grant, training award or capacity funding) from their programme names. They were added in count-based analyses, and their amounts were imputed from priced grants of the same funder, funder country or award type, at the 25th, 50th and 75th percentiles. Second, yearly records of the same project were merged. Third, we varied the timing of funding (spread over grant years), the income group (at grant start), the definition of the Global South (OECD Development Assistance Committee list), the treatment of NIAID (excluded), the dollar basis (constant 2023 dollars) and the topic assignment (topic probabilities from a refitted model). Finally, the gender model was re-estimated per year of grant, without funders in Japan and China, where name-based inference is less reliable, and without training awards.

### 2.5 Reproducibility

All analyses were run in Python (pandas, statsmodels, scikit-learn, gensim, spaCy). Seven notebooks rebuild every table and figure, and every number quoted in this paper, from the raw export with one script. A clean rebuild reproduced all 43 output tables byte for byte. The code is openly available (see Data availability).

## 3. Results

### 3.1 Growing, but small

The 802 grants came from 105 funders in 25 countries and went to 525 institutions in 39 countries. Together they recorded USD 318.6 million over 25 years, or about USD 13 million a year. The median grant was USD 180,000 (interquartile range 40,400–428,116).

Funding grew fast from this small base. Grants grew by 12.2% a year (95% CI 10.0–14.5) and recorded funding by 16.6% a year (12.6–20.7) over 1999–2022 (Figure 1, Supplementary Table S2). In constant 2023 dollars, funding grew by 14.1% a year (10.2–18.2). Two thirds of all funding (USD 213.0 million, 502 grants) started in 2016–2023, against USD 29.1 million (102 grants) in 1999–2008 (Table 1). Yearly totals were uneven, with peaks in 2009 and 2018, and reached about USD 30–39 million a year in 2018– 2022.

**Figure 1.**
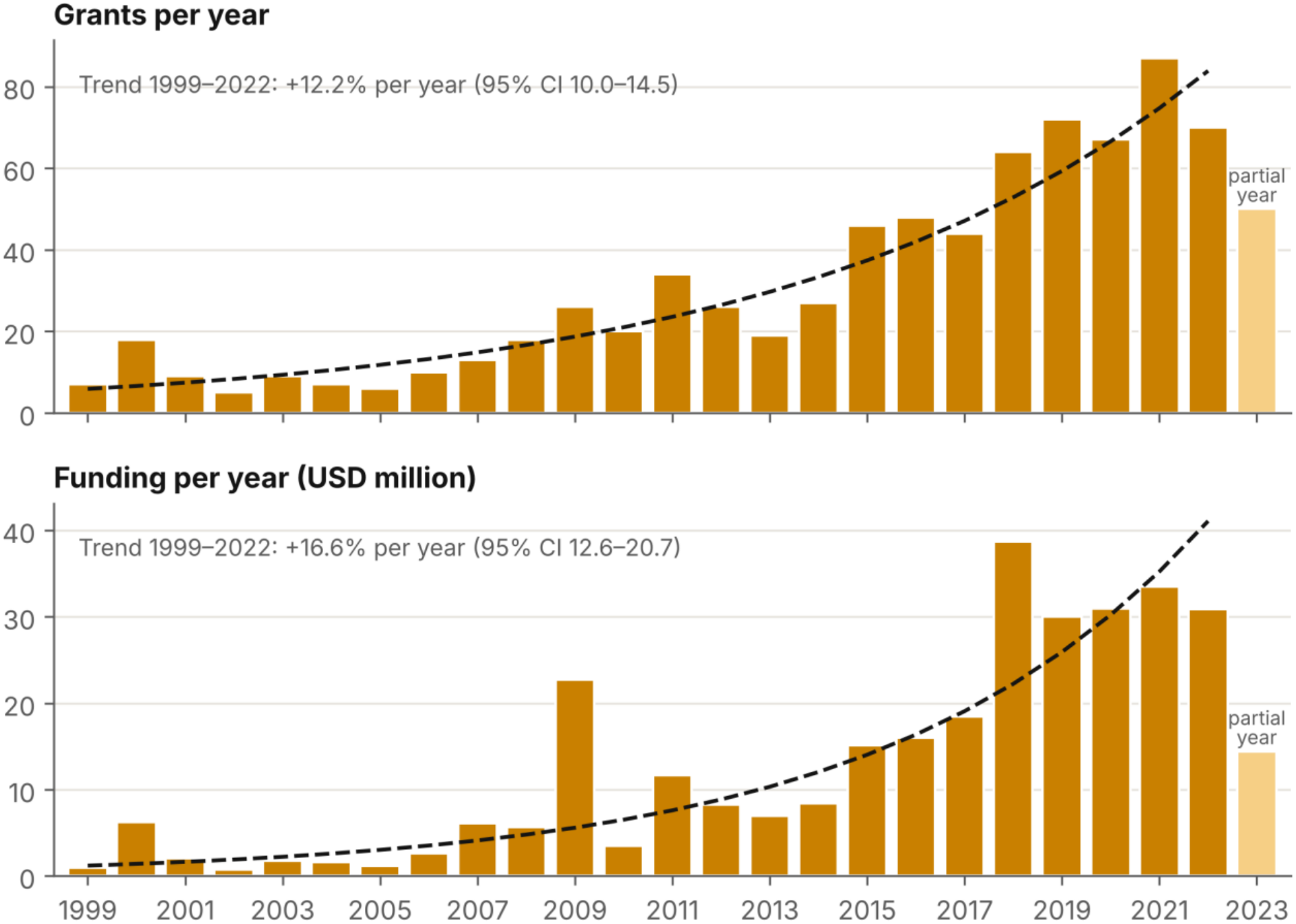
Grants grew about 12¾ a year and funding about 17¾ a year, 1999-2022. Grants (top) and recorded funding (bottom) per year, with log-linear trends for 1999–2022. 2023 is a partial year.

**Table 1.** Grants and recorded funding by period of start year.

| Period | Grants | Total (USD M) | Median (USD) | Interquartile range (USD) |
|---|---|---|---|---|
| 1999–2008 | 102 | 29.1 | 124,007 | 28,188–368,145 |
| 2009–2015 | 198 | 76.6 | 140,656 | 48,250–410,679 |
| 2016–2023 | 502 | 213.0 | 211,446 | 40,673–444,375 |
| All years | 802 | 318.6 | 180,000 | 40,400–428,116 |

### 3.2 Concentrated in one country and one institute

US funders provided 47.7% of recorded funding, and one institute, NIAID, provided 21.0% (Table 2, Figure 5). NIAID’s share rose across the three periods, from 13.6% in 1999–2008 to 17.4% in 2009–2015 and 23.3% in 2016–2023. The next largest funders were the US National Cancer Institute (4.9%), the European Commission (4.6%) and the Swiss National Science Foundation (3.7%). By number of grants, the leaders were the Japan Society for the Promotion of Science (105 grants), NIAID (81) and the National Natural Science Foundation of China (79), which fund many smaller grants.

**Table 2.** Ten largest funders and recipient institutions by recorded funding. Recipients use fractional counting.

| Funder | Country | Grants | USD M | Share (%) |
|---|---|---|---|---|
| National Institute of Allergy and Infectious Diseases | United States | 81 | 66.9 | 21.0 |
| National Cancer Institute | United States | 12 | 15.5 | 4.9 |
| European Commission | European Union | 19 | 14.6 | 4.6 |
| Swiss National Science Foundation | Switzerland | 26 | 11.8 | 3.7 |
| Biotechnology and Biological Sciences Research Council | United Kingdom | 24 | 11.2 | 3.5 |
| Federal Ministry of Education and Research | Germany | 10 | 10.1 | 3.2 |
| Congressionally Directed Medical Research Programs | United States | 21 | 9.2 | 2.9 |
| National Institute of General Medical Sciences | United States | 11 | 9.1 | 2.9 |
| Ministry of Industry and Trade | Czechia | 5 | 8.5 | 2.7 |
| Canadian Institutes of Health Research | Canada | 15 | 7.8 | 2.5 |
| Intralytix (United States) | 9 | 8.46 |  |  |
| Division of Basic Sciences, NCI (United States) | 1 | 7.22 |  |  |
| Purdue University West Lafayette (United States) | 3 | 6.90 |  |  |
| Dyntec (Czechia) | 2 | 6.75 |  |  |
| University of Montana (United States) | 2 | 5.76 |  |  |
| Fraunhofer Society (Germany) | 1 | 5.24 |  |  |
| University of Toronto (Canada) | 9 | 5.17 |  |  |
| Stanford University (United States) | 7 | 4.83 |  |  |
| MB Pharma (Czechia) | 2 | 4.71 |  |  |
| Kagoshima University (Japan) | 10 | 4.46 |  |  |

Funding was highly concentrated by country: the HHI across funder countries was 2,501, above the usual threshold of 2,500. Without NIAID, it fell to 1,508 and the US share to 33.8%, while growth was almost unchanged (16.2% a year; Supplementary Table S27). Across recipient institutions, the top 10% received 53.4% of funding (Gini 0.68; Figure 5b). The largest single recipient, Intralytix in the United States, received 2.7%.

### 3.3 Unequal geography

Funding stayed almost entirely within high-income countries. Funders in high-income countries provided 98.3% of recorded funding, and institutions in high-income countries received 97.8% (Figure 4, Supplementary Table S4). Low- and lower-middle-income countries received 0.33%. Global South funders provided USD 5.6 million (1.7%), and Global South recipients received USD 6.7 million (2.1%), 73% of it in China (Supplementary Table S11).

Most funding also stayed within each funder’s own country. Grants whose recipients were all in the funder’s country accounted for 88.7% of funding (Supplementary Table S12). The European Commission was the largest cross-border funder, with 19 grants and USD 14.6 million. Collaboration across the income divide was rare. Of the 135 grants with more than one recipient institution, 39 involved more than one country, and only 7 linked Global North and Global South recipients (Supplementary Table S26).

### 3.4 What, and who, gets funded

Four research topics emerged: infectious disease control (280 grants; 35.9% of funding), bacterial genetics and mechanisms (237; 28.8%), antibiotic resistance strategies (151; 22.4%), and immunotherapy and targeted treatments (133; 12.9%) (Figure 2, Supplementary Table S37). Their balance shifted over time. Antibiotic-resistance research received 3% of funding in 1999–2005 and 5% in 2006–2010, then about a quarter from 2011 onward (28%, 25% and 27% in the three later periods). Immunotherapy fell from half of the small early funding to about 10%.

**Figure 2.**
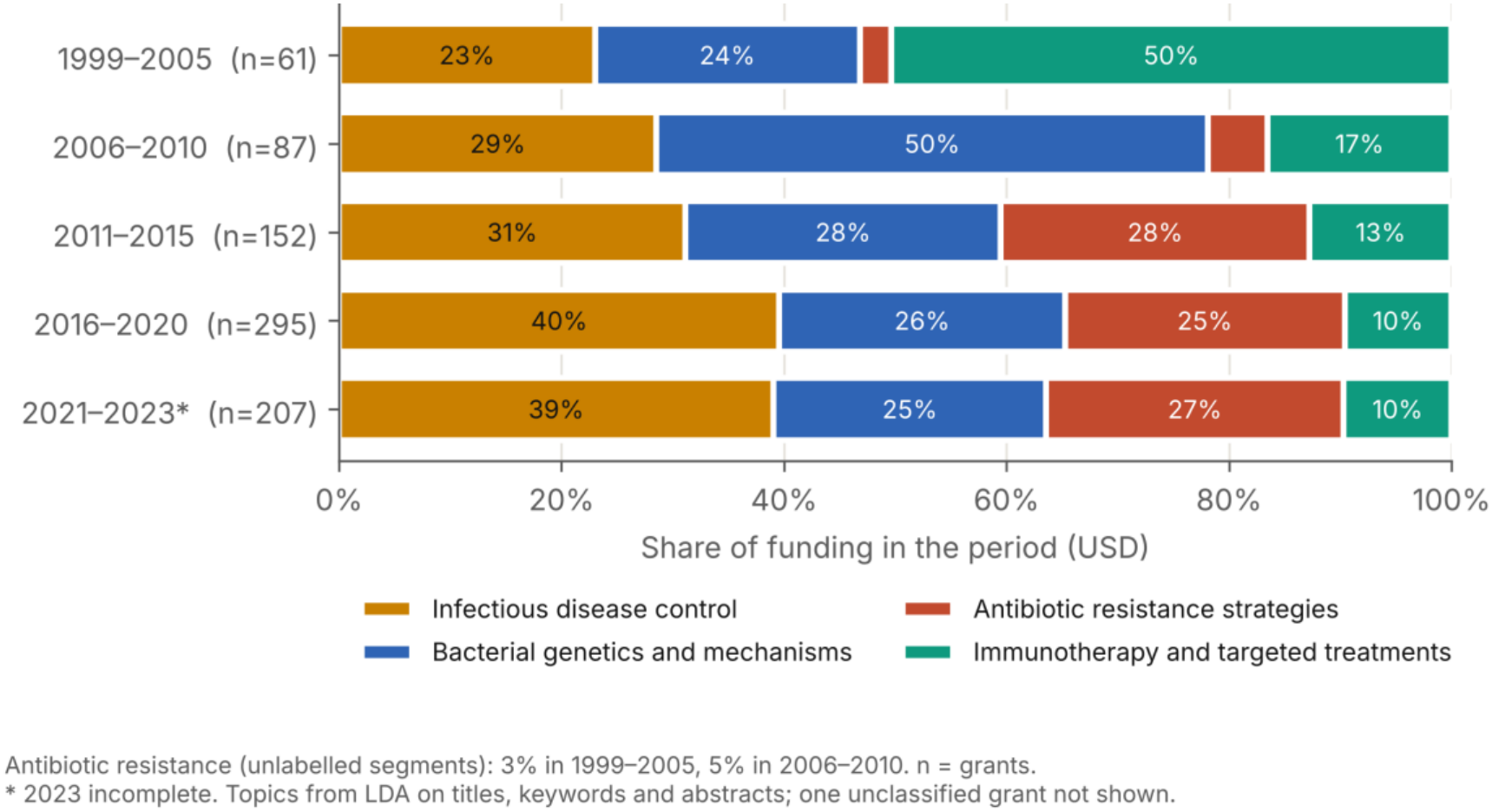
Antibiotic-resistance research rose from 3% to about a quarter of funding. Share of recorded funding by LDA research topic in five periods. n = grants per period.

Funders specialised. NIAID placed 58% of its grants in antibiotic resistance, the National Natural Science Foundation of China 44% in immunotherapy, and the US National Cancer Institute 83% in immunotherapy (Supplementary Figure S3).

Of 1,073 named researchers, 373 were inferred as women and 696 as men; women made up about a third of named researchers each year from 2011 to 2022 (Supplementary Figure S6). Male-only teams held 446 grants (USD 179.0 million), female-only teams 201 (USD 57.4 million) and mixed teams 95 (USD 41.6 million). A further 60 grants (USD 40.7 million) had no researcher of known gender (Table 3, Figure 3). After adjustment for team size, grant length, start year, funder and topic, female-only teams received grants 19% smaller than male-only teams (95% CI −32 to −2; p = 0.026). Per year of grant, the gap was 22% (−35 to −6). Mixed teams did not differ from male-only teams after adjustment (−7%, p = 0.61).

**Figure 3.**
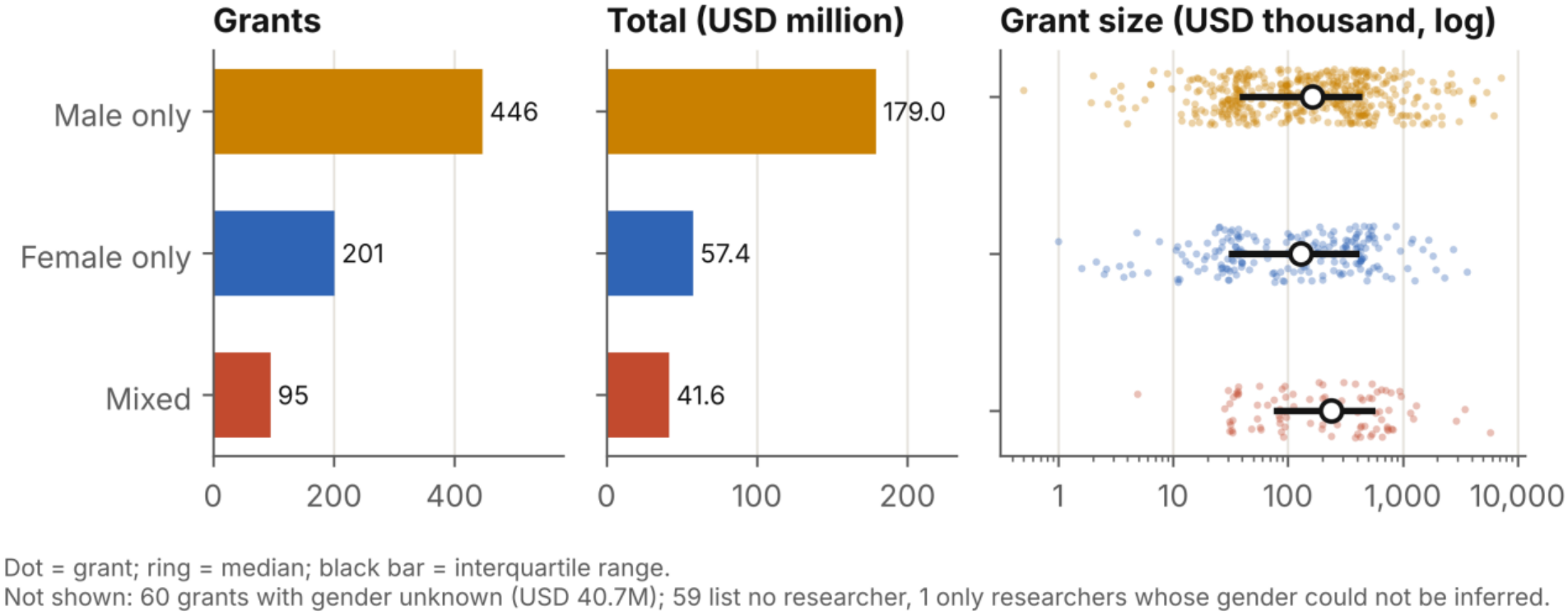
Female-only teams receive fewer and smaller grants. Grants, total funding and grant size by gender composition of the research team. Dots are grants; rings are medians; bars are interquartile ranges.

**Figure 4.**
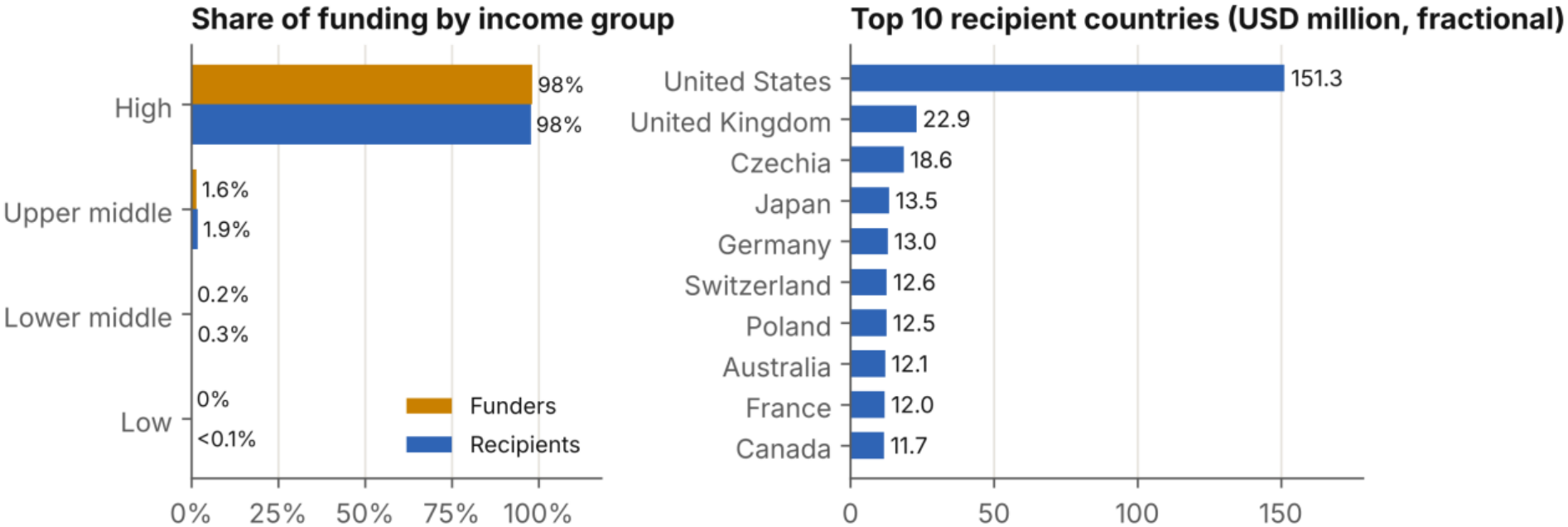
Low- and lower-middle-income countries received 0.3% of funding. Share of recorded funding by World Bank income group for funders and recipients (left), and the ten largest recipient countries (right, fractional counting).

**Figure 5.**
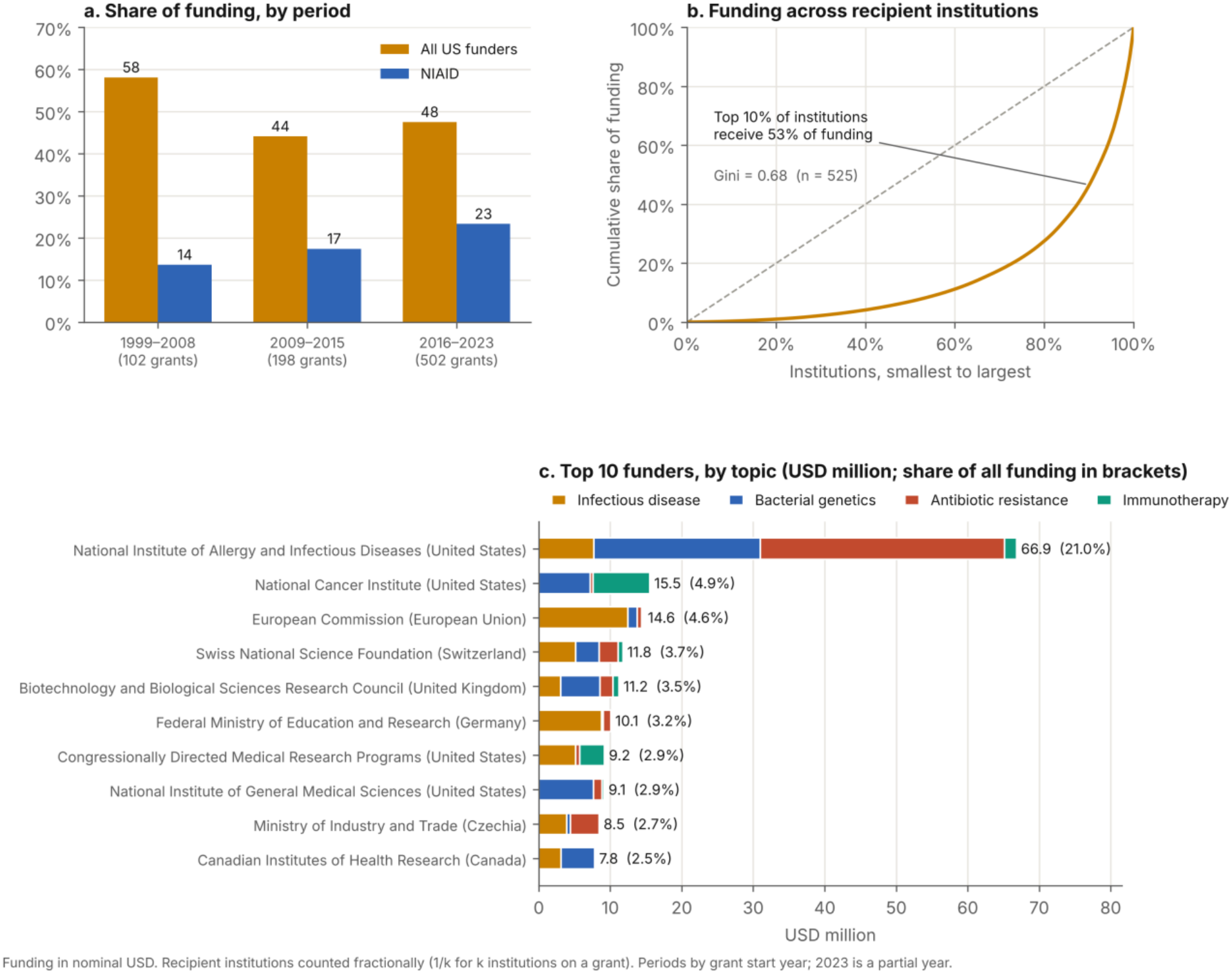
Funding is concentrated in one country, one institute and a few institutions. Concentration: (a) US and NIAID shares by period; (b) Lorenz curve across recipient institutions; (c) the ten largest funders, by topic.

**Figure 6.**
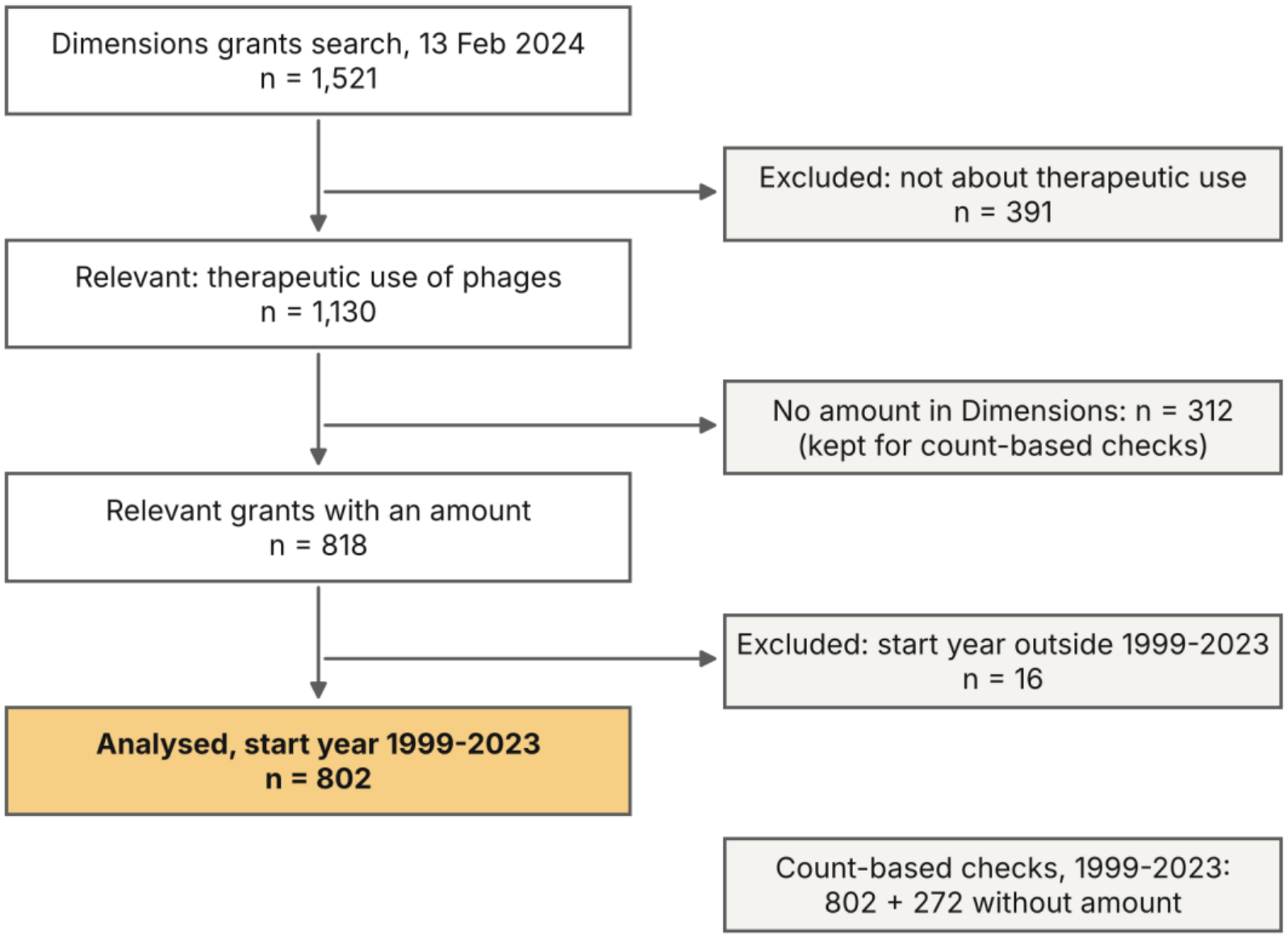
From the Dimensions search to the analysed grants (repository Figure S11).

**Table 3.**
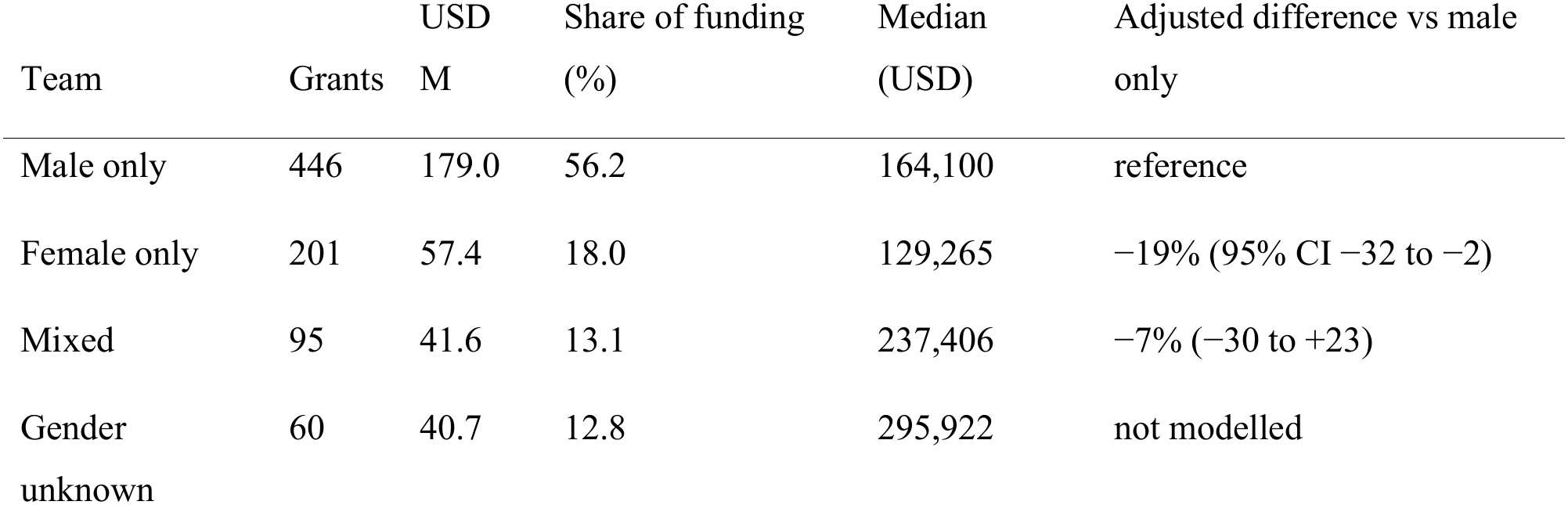
Grants by gender composition of the research team. Adjusted difference from a regression of log(amount) on team group, team size, grant length, start year, funder and topic.

| Team | Grants | USD M | Share of funding (%) | Median (USD) | Adjusted difference vs male only |
|---|---|---|---|---|---|
| Male only | 446 | 179.0 | 56.2 | 164,100 | reference |
| Female only | 201 | 57.4 | 18.0 | 129,265 | −19% (95% CI −32 to −2) |
| Mixed | 95 | 41.6 | 13.1 | 237,406 | −7% (−30 to +23) |
| Gender unknown | 60 | 40.7 | 12.8 | 295,922 | not modelled |

Most researchers appeared once. Of the 1,073 named researchers, 83.9% held a single grant. Among the 525 recipient institutions, 61% appeared in only one year (Supplementary Figures S8–S9).

### 3.5 Robustness

The 272 relevant grants without an amount were not a random subset. Training awards made up 43% of them (studentships, scholarships and theses), against 8% of priced grants, and most came from funders in Brazil, Russia and Belgium (Supplementary Table S30). Counting grants rather than dollars, the Global South received 17.5% of the 1,074 relevant grants, against 11.8% of the 802 priced ones (Supplementary Table S31). With imputed amounts, its share of funding rose from 2.1% to 2.6–5.5% (Supplementary Table S32).

The other headline results held. Low- and lower-middle-income countries stayed at or below 0.6% of funding and 1.4% of grants. Growth stayed at 16–17% a year, the US share at 41–48% and NIAID’s at 17–21%. The gender gap held in every subset (−19% to −22%). Merging yearly records into 771 projects changed no result by more than one point. Overall, 3 of 40 checks changed a conclusion: the two count-based Global South measures, and the year-based “transient” label, which falls from 82% to 3.5% when every year of a grant counts (Supplementary Figure S12). For this reason we report retention as the share of researchers holding a single grant, which does not depend on that choice.

## 4. Discussion

### 4.1 Main findings

Over 25 years, recorded funding for phage therapy research grew by about 17% a year. It remained small, about USD 13 million a year on average. It is concentrated in one country and one institute: US funders provide nearly half of it and NIAID a fifth, a share that is rising. It rarely leaves high-income countries, and low- and lower-middle-income countries receive a third of one percent. Antibiotic resistance has become a central topic. Female-only teams receive smaller grants, and most researchers hold a single grant.

### 4.2 A mismatch between burden and funding

The countries where AMR kills most people are almost absent from this funding. In 2019, the highest death rates attributable to AMR were in sub-Saharan Africa and South Asia [1]. In our data, low- and lower-middle-income countries received 0.33% of recorded funding and 1.4% of grants, and Uganda and Tanzania were the only recipient countries in sub-Saharan Africa outside South Africa. This pattern is not unique to phage therapy: investment in infectious disease research also follows the location of funders rather than of disease [10]. It matters more for phages than for most drugs, however. Phages are specific to bacterial strains, so phage banks and cocktails must be built from local isolates, and production close to patients is part of the appeal of the approach [3,4]. A field funded almost entirely in high-income countries may build its libraries and its evidence around their pathogens.

The Global South is not inactive. Counting grants rather than dollars, it holds 17.5% of relevant grants, mainly because funders in Brazil report no amounts to Dimensions; Russian funders do the same. Data visibility is therefore part of the inequity. Global funding trackers, and analyses such as ours, can only measure what funders report, and they under-count national investment in exactly the regions that most need to be seen.

### 4.3 Fragility

A field that draws nearly half its funding from one country, and a fifth from one institute, depends on that institute’s priorities and budget. NIAID’s support has been decisive: it funds 58% of its phage grants in antibiotic resistance and, in 2026, launched three Centers for Accelerating Phage Therapy [7]. The same dependence is a risk. By April 2026, the US Department of Health and Human Services had terminated 1,392 NIH awards before completion, against fewer than six from 2012 to January 2025 [18]. Our sensitivity analysis shows that growth does not hinge on NIAID alone (16.2% a year without it), but the field’s centre of gravity does.

### 4.4 A thin pipeline

Most researchers (84%) appear on a single grant, and most institutions in a single year. Clinical trials, manufacturing to good manufacturing practice and regulatory dossiers need teams that stay in the field for years. A workforce that turns over this quickly may slow translation more than the level of funding does. Longer awards, programme grants and fellowships that keep phage researchers in the field would address this directly.

### 4.5 Gender

Women made up about a third of named researchers, and female-only teams received grants about a fifth smaller than male-only teams, after accounting for funder, grant length and topic. This matches findings in other fields [19,20]. Mixed teams’ higher average grants disappeared after the same adjustment, so our data do not support a funding advantage for mixed teams; the unadjusted difference was accounted for by funder and grant length. Name-based inference is binary and less reliable for some languages, but the gap held when funders in Japan and China were excluded.

### 4.6 Implications

Four steps follow from these results:

1. **North–South partnerships and shared funding pools**, so that phage research and trials include the countries with the highest AMR burden. Only 7 of 802 grants linked the two today.
2. **Funding for phage banks and production capacity in low- and middle-income countries**, since phage therapy depends on local isolates and local manufacture.
3. **Tracking phage therapy separately in AMR funding dashboards**, and asking national funders to report award amounts, so that investment outside the major Western funders becomes visible.
4. **Monitoring gender in phage therapy funding**, including grant size and not only grant counts.

### 4.7 Limitations

First, dollar results cover the 802 grants with a recorded amount. Of the 1,074 relevant grants that started in 1999–2023, 272 had no amount; 43% of these were training awards, and most came from funders in Brazil, Russia and Belgium. Count-based and imputation analyses show which results depend on this gap. Second, Dimensions covers NIH, European and Japanese funders well but many national funders poorly, including those of Georgia, home of the Eliava Institute, and Russia, both long-standing centres of phage therapy. Third, private, philanthropic and venture funding is largely out of scope, although companies such as Intralytix appear as grant recipients. Fourth, 2023 is incomplete because of indexing lag, and amounts are nominal US dollars converted by Dimensions; constant-dollar growth was similar. Fifth, gender was inferred from first names and is binary. Sixth, LDA topics are approximate: assignments varied between model runs, so we report topic trends rather than precise shares. Seventh, all results are descriptive associations; in particular, the gender model adjusts for observed differences between grants but cannot identify why they differ. Finally, we do not link grants to outputs such as publications, patents or trials; that link is the natural next step from our patent study [11].

## 5. Conclusion

Recorded funding for phage therapy research grew by about 17% a year over 25 years, but from a small base and on a narrow footing. Nearly half comes from US funders and a fifth from one institute, and almost none reaches the low- and lower-middle-income countries that carry the heaviest burden of antimicrobial resistance. As phage therapy moves toward regulated clinical use, broadening who funds it, where the money goes and who holds the grants will shape whom it serves. Making every funder’s investment visible is the first step.

## Supporting information

Supplementary_file_3_BIBLIO_checklist

## Declarations

### Ethics statement

This study analysed records of research grants. It did not involve contact with people, interventions or clinical data. Investigator names are listed publicly in the grant records; they were used only to infer team gender, and no individual is identified in the results or in the public outputs.

### Disclaimer

The contributions of the NIH author(s) are considered Works of the United States Government. The findings and conclusions presented in this paper are those of the author(s) and do not necessarily reflect the views of the NIH or the U.S. Department of Health and Human Services.

### Data availability

All code, the country classification table, and every aggregate table and figure are openly available at https://github.com/descartesmbogning/phage-therapy-funding-complete. Grant-level data cannot be redistributed under the Dimensions terms of use. They can be rebuilt with the published query and a Dimensions subscription, after which the pipeline reproduces every output. Researcher names and inferred gender are not shared.

### Code availability

The analysis code is released under the MIT licence in the same repository.

### Reporting guideline

This report follows the BIBLIO guideline for bibliometric studies [21] where its items apply to grant records. The completed checklist is provided as a supplementary file.

### Use of AI tools

An AI assistant (Claude, Anthropic) was used to help review the analysis code and to edit the text. The authors checked all outputs and take full responsibility for the content.

### Trial registration

Not applicable.

## Supplementary material

**Supplementary file 1 (PDF).** Supplementary Methods (full search query, screening, classifications, topic model, sensitivity analyses), Supplementary Figures S1–S12 and Supplementary Tables S1–S38. Supplementary Figure S11 is the screening flow shown as Figure 6; Table 3 combines the repository tables Table4_gender and S3_gender_tests; Supplementary Tables S37 and S38 are the repository tables Table3_topics and Table5_income_groups.

**Supplementary file 2 (XLSX).** All supplementary tables, one sheet per table.

**Supplementary file 3 (XLSX).** BIBLIO reporting checklist.

