## Supplementary_file_1_methods_figures_tables for "Twenty-five years of phage therapy funding: rapid growth, narrow base"

Every table and figure here is generated by the code in the open repository (https://github.com/descartesmbogning/phage-therapy-funding-complete); the repository name of each item is given in brackets. Amounts are recorded US dollars as converted by Dimensions; shares of funding are shares of recorded funding (802 grants, USD 318.6 million) unless stated.

### Contents


- Supplementary Methods
- Supplementary Figures S1–S12
- Supplementary Tables S1–S38

### Supplementary Methods

#### 1. Search (Dimensions, 13 Feb 2024, no date limit)

```
(title:(Bacteriophage OR Bacteriophages OR phage OR phages OR "phago virus" OR "phagovirus" OR siphophage* OR Microbacteriophage* OR
Mycobacteriophage* OR Mycobacteriophage OR Microbacteriophage OR multiphage* OR multiphage OR "multi-phage*" OR "phage-based" OR
"bacteriophage-based" OR phageome OR phagotherapy OR "Monophage Therapy" OR "Polyphage" OR Coliphage OR prophage OR staphylophage OR
phageome OR PHAGOTHERAPY)) AND ((abstract:(therap* OR Therapy OR THERAPEUTIC OR treat* OR Treatment OR antibacterial OR
antibacteria* OR "anti-bacterial" OR antimicrobial OR cocktail* OR cocktail OR prophylax* OR Prophylaxis OR Antifungal OR Antiviral
OR PHAGOTHERAPY)) OR (title:(therap* OR Therapy OR THERAPEUTIC OR treat* OR Treatment OR antibacterial OR antibacteria* OR "anti-
bacterial" OR antimicrobial OR cocktail* OR cocktail OR prophylax* OR Prophylaxis OR Antifungal OR Antiviral OR PHAGOTHERAPY)))
```

#### 2. Screening (notebook 06, Figure S11, Table S29)

| Step | Removed | Left |
|---|---|---|
| Dimensions search | | 1,521 |
| Not about therapeutic use of phages (titles and abstracts) | 391 | 1,130 |
| No funding amount in Dimensions | 312 | 818 |
| Start year outside 1999–2023 | 16 | 802 |

Dollar results use the 802 grants with a recorded amount. The 272 relevant grants without an amount that started in 1999–2023 are used in the robustness checks (section 7). The counts are read from the screening columns of the export (`includes ?`, `funding?`), so the flow diagram is generated, not typed.

#### 3. Enrichment

- **Institutions.** Harmonised with GRID/ROR in Dimensions, then checked by hand. Missing co-recipients were added.
- **Countries.** Funder and recipient countries get a World Bank income group (fiscal year 2024) and a Global North/South label (World Population Review 2024) from one table, `data/reference/country_classification.csv`.
  - Georgia, Russia, Ukraine, Singapore and Qatar are labelled differently by different sources; they keep the source label and are flagged in the table.
  - The European Commission and the European Research Council are coded as funder country "European Union".
- **Gender.** Researcher gender is inferred from first names with Gender API; ambiguous names were checked by hand.

#### 4. Topics (notebook 02)

1. **Text.** Title, abstract and keywords, using the English translations.
2. **Stopwords.** WordCloud list, NLTK stopwords in all languages, and "amp".
3. **Duplicates.** Exact duplicate texts are fitted once; each duplicate takes its twin's topic.
4. **Lemmas.** Nouns, adjectives and verbs only (spaCy `en_core_web_md`).
5. **Term counts.** Tokens longer than 2 characters, `min_df=50`, `max_df=0.75`.
6. **LDA.** scikit-learn, online learning, `learning_offset=50`, seed 111.
7. **Choice of k.** Mean c_v coherence over the top 30 words, k = 2–12; highest at k = 5 (Figure S1).
8. **Labels.** Topics were labelled from their top words and the abstracts that fit each topic best:
   - 0: Bacterial genetics and mechanisms
   - 1: Antibiotic resistance strategies
   - 2: Immunotherapy and targeted treatments
   - 3: one grant, reported as Unclassified
   - 4: Infectious disease control
9. **Stability.** Refitting with five other seeds keeps 51–68% of grants in the same topic (Table S9), so results are reported for topic totals, not single grants.
10. **Stored assignment.** Model fits vary slightly with library versions, so the topic of each grant used in the paper is stored (`data/raw/lda_topic_assignment.pkl`). Notebook 02 refits the model, compares the refit with the stored assignment, and saves the refit's topic probabilities for the robustness checks.

#### 5. Analysis (notebook 03)

- **Growth.** Log-linear fit of yearly counts and USD totals over 1999–2022, reported as % per year with a 95% CI. 2023 is excluded from the fit because of indexing lag.
- **Fractional counting.** A grant with *k* recipient institutions gives 1/*k* of its amount to each one, and through it to that institution's country and income group. Each grant has one funder.

- **Domestic vs cross-border.** A grant is domestic when every recipient institution is in the funder's country (Table S12): 88.7% of funding. Splitting multi-country grants fractionally instead gives 91.4%, because 30 of the 65 cross-border grants also include a recipient in the funder's country.
- **Gender.**
    - Exclusive groups by the genders of named researchers: male only, female only, mixed, and gender unknown (59 grants list no researcher; 1 lists researchers whose gender could not be inferred).
    - Groups are compared with Mann–Whitney tests.
    - Main model: OLS of log(amount) on group, log team size, log grant length (years), start year, funder (funders with fewer than 10 grants pooled) and topic, with HC1 robust errors. Table S3 also reports the amount per year of grant and a simple model (team size, US funder, start year).
- **Engagement types** (as in the patent study). In each year an entity is active it is a **Newcomer** (not active before, active later, or first seen in 2023), **Continuant** (active before and after), **Terminator** (active before, not after) or **Transient** (active in one year only). The summary uses each entity's last label. Because the shares depend on whether a grant counts in its start year only or in every year it runs (Table S7), the share of researchers who held a single grant is the main retention measure.

#### 6. Concentration, collaboration and sensitivity (notebook 05)

- **Concentration.**
    - Herfindahl–Hirschman index (HHI): the sum of squared percentage shares, 0–10,000, for funders, funder countries and recipient countries (fractional counting). By convention, above 2,500 is highly concentrated and 1,500–2,500 moderately concentrated.
    - Gini coefficient and the share received by the top 10% of recipient institutions (fractional counting). With 525 institutions, the top 10% is the 52 largest.
    - Shares from US funders and from NIAID, for 1999–2008, 2009–2015 and 2016–2023.
- **Collaboration.** Counts only: grants with more than one recipient institution or country, grants linking Global North and Global South recipients, co-investigator pairs.
- **Without NIAID.** Growth, the US share and the HHIs re-estimated with NIAID grants removed.
- **Constant dollars.** Amounts converted to 2023 USD with the annual average CPI-U (all items, U.S. city average, 1982–84 = 100), by grant start year (`data/reference/cpi_u_annual.csv`: 1999–2021 from the BLS CPI-U historical table; 2022 = 292.655 and 2023 = 304.702 from BLS data as published by the Idaho Department of Labor). Dimensions had already converted other currencies to USD, so only US inflation is removed.

#### 7. Robustness (notebook 06)

- **Award types.** Each record is classed as a research grant, a training award (studentship, scholarship, fellowship, thesis, graduate-programme record) or capacity funding (USDA formula funds) from its programme name and source link (`robustness.award_type`). 43% of the grants without an amount are training awards, against 8% of grants with one (Table S30).
- **Grants without an amount.** The 272 relevant grants with no amount enter three checks:
    - count-based shares, each grant counted once and split equally across its recipient countries (Table S31);
    - imputation: each missing amount takes the median (central), 25th percentile (low) or 75th percentile (high) of priced grants from the same funder (at least 3), else the same funder country and award type, else the same award type. Training awards are 0 in the low and central scenarios and the 25th percentile of priced training awards in the high scenario (Table S32);
    - recovery: amounts found by hand on funder websites, saved in `data/raw/recovered_amounts.csv`, are used when present.
- **Projects.** Records with the same funder and the same English (translated) title whose years follow on are merged into one project, with amounts summed (Table S33).
- **Other definitions.**
    - Amounts spread evenly over grant years.
    - Income group at grant start: China lower-middle income until 2009 and Thailand until 2010 (to be checked against the World Bank historical classification, OGHIST).
    - OECD DAC list of ODA recipients (2024–25) instead of Global South.
    - Engagement: share of researchers with one active year among those first seen 2001–2019, and share with a single grant.
    - Topic shares from the refitted model's topic probabilities (Table S35).
    - Gender model without funders in Japan and China (name inference is less reliable there) and without training awards (Table S34).
- **Validation samples and cross-checks.** The notebook writes private samples for a second screener (15% of 1,521, stratified), hand-coding of topics (100 grants) and checking of inferred gender (100 names, blind), and reports Cohen's kappa once they are filled. Public templates hold yearly NIH totals (for comparison with NIH RePORTER) and World Bank price level ratios (for a PPP check).
- **Summary.** Figure S12 and Table S36 show each headline under each check, with the rule used to judge whether its conclusion holds.

#### 8. Paper numbers (notebook 07)

`src/paper_numbers.py` recomputes every number quoted in the manuscript and records its source (`outputs/tables/paper_numbers.csv`). Notebook 07 checks the manuscript text against it in both directions.

### Supplementary Figures

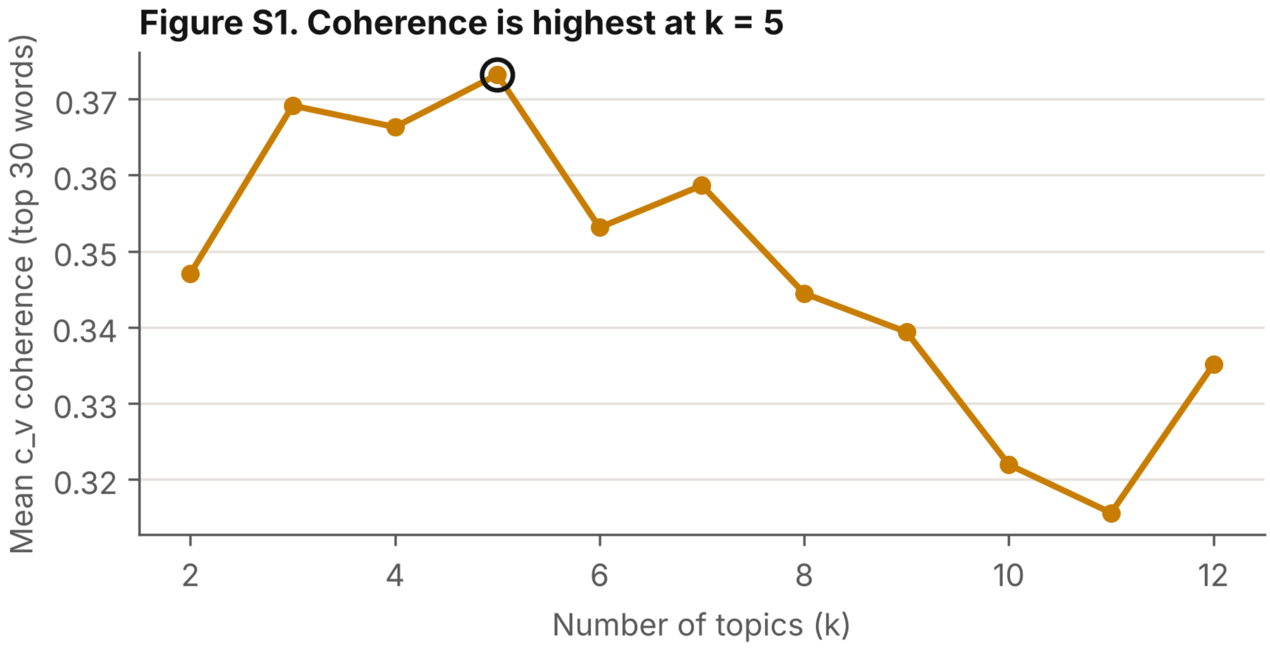


**Figure S1.** Topic coherence (c_v) by number of topics; k = 5 was chosen. [FigS1_topic_coherence]

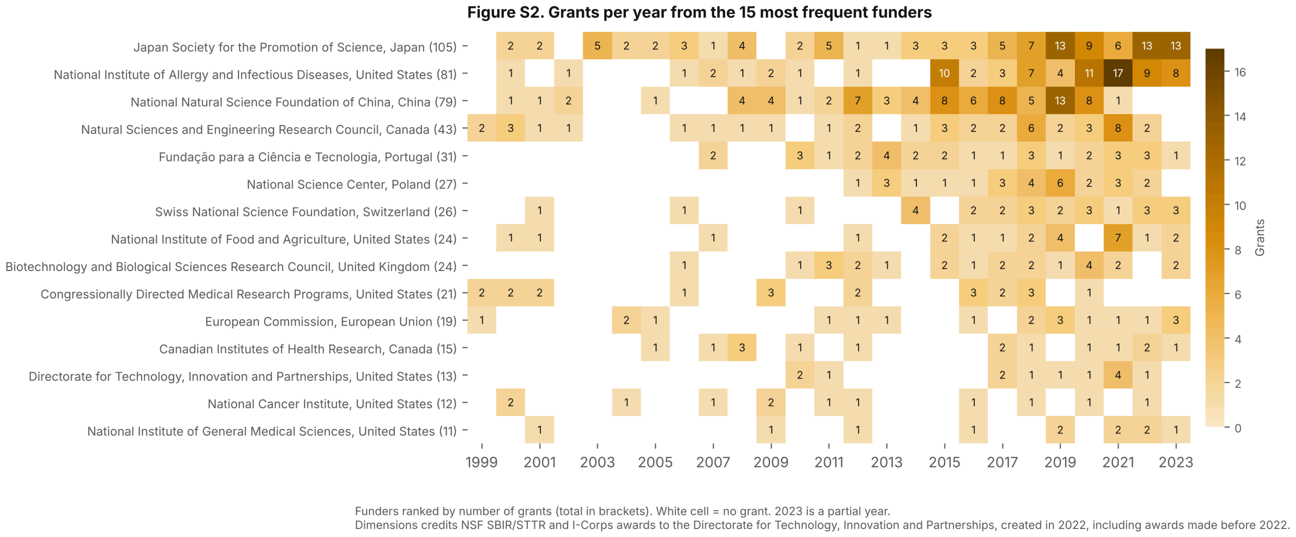


**Figure S2.** Grants per year for the 15 funders with the most grants. [FigS2_funders_by_year]

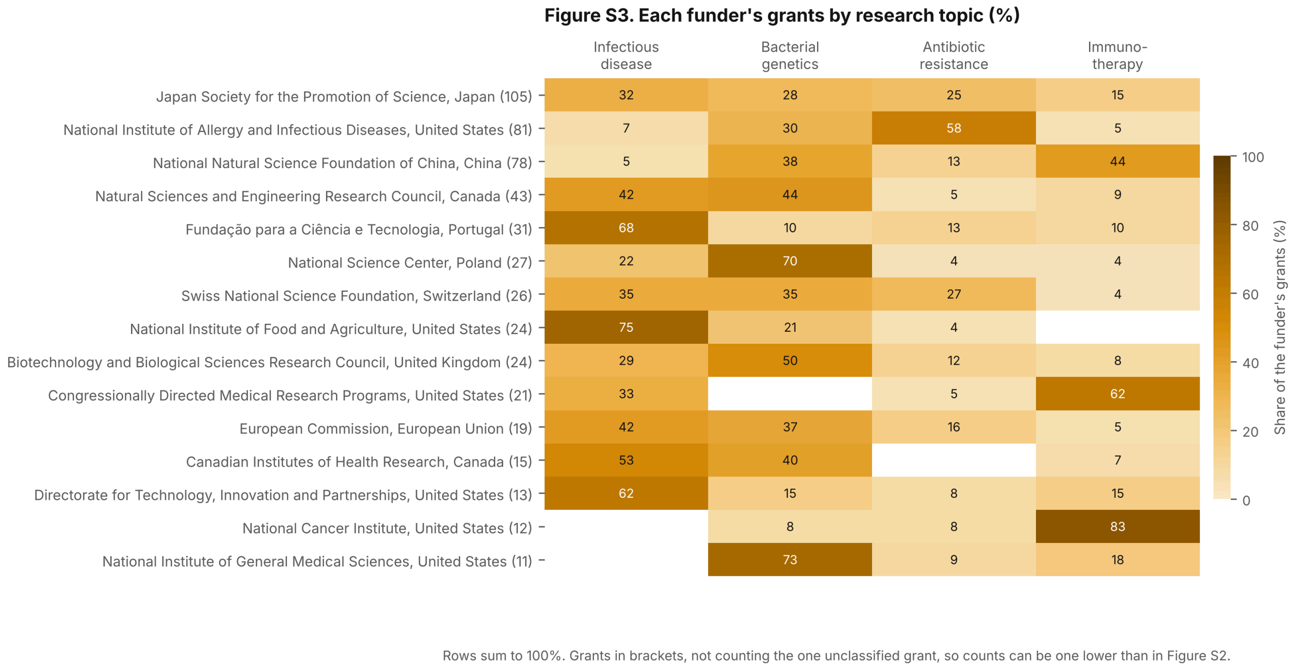


**Figure S3.** Each funder's grants by research topic. [FigS3_topic_by_funder]

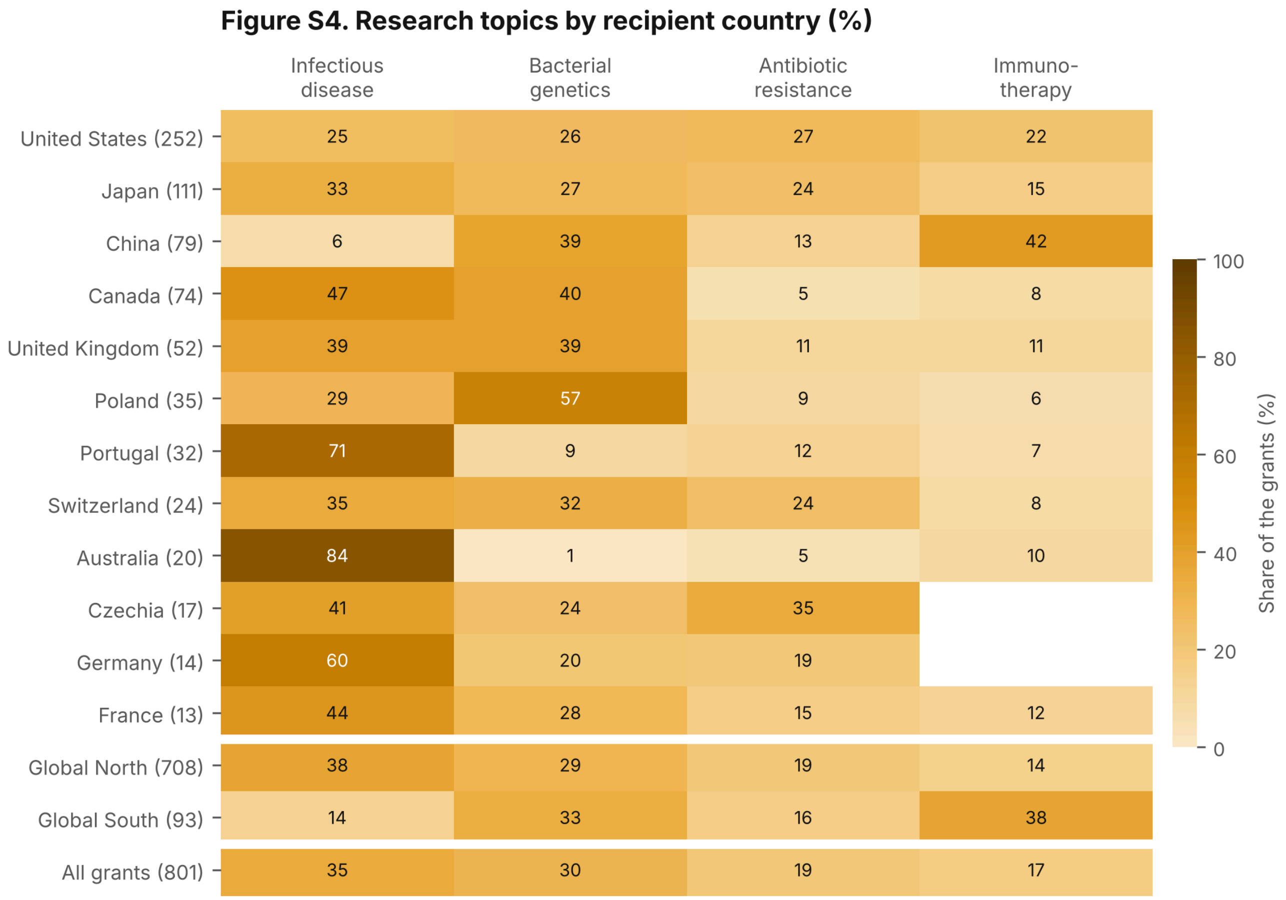


Fractional counting: a grant with k recipient institutions counts 1/k for each one's country.
12 countries with most grants, then all Global North and Global South recipients. Rows sum to 100%.

**Figure S4.** Research topics by recipient country. [FigS4_topic_by_recipient]

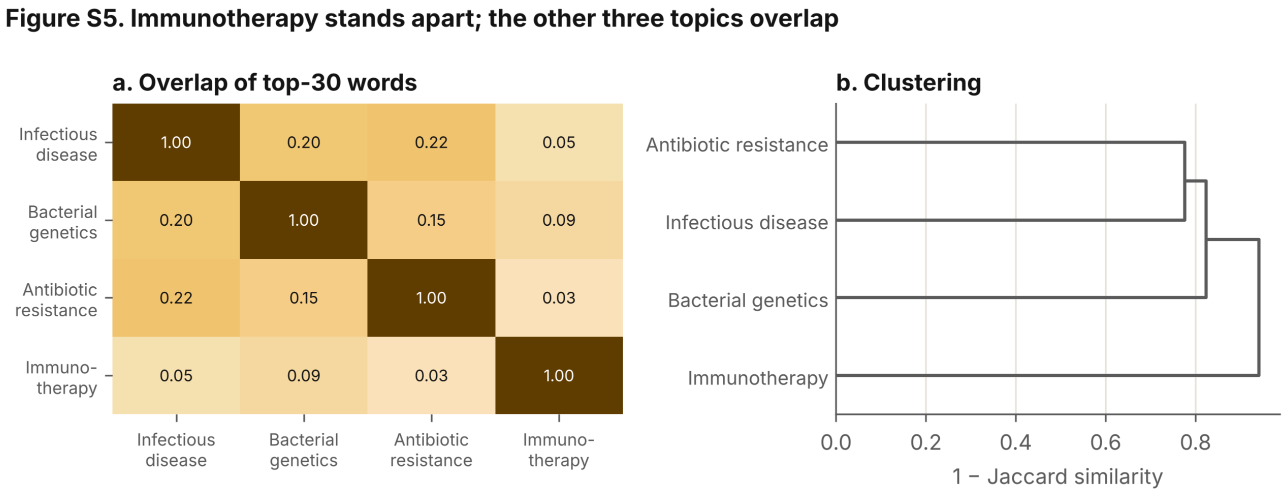


Top-30 words per topic by mean TF-IDF of its grants (words in over half of all grants removed), LDA topic assignment used in the paper. Word lists: Table S17.

**Figure S5.** Topic similarity and distinctive words. [FigS5_topic_similarity]

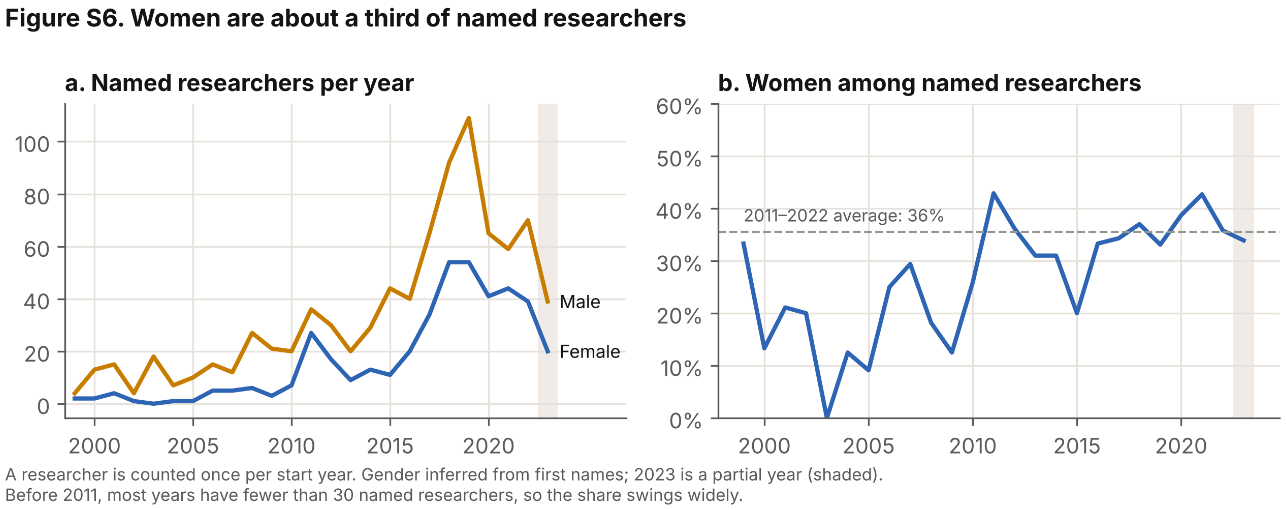


**Figure S6.** Named researchers by inferred gender per year. [FigS6_researchers_by_gender]

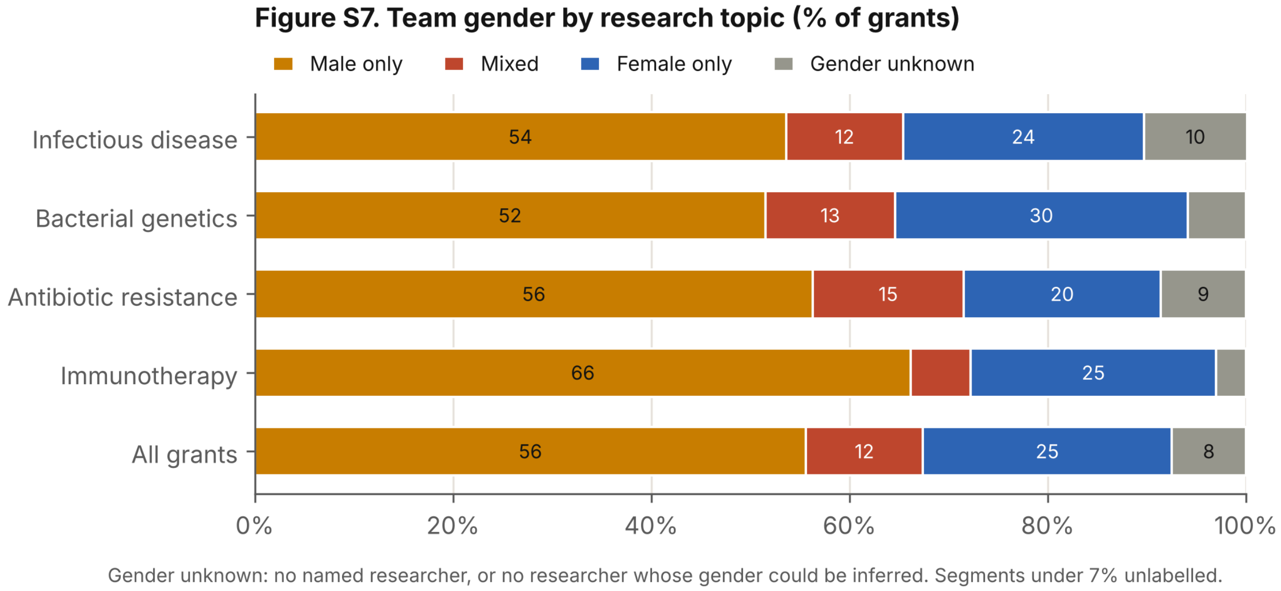


**Figure S7.** Team gender composition by research topic. [FigS7_team_gender_by_topic]

### Figure S8. Most researchers appear in one year only (Transient)

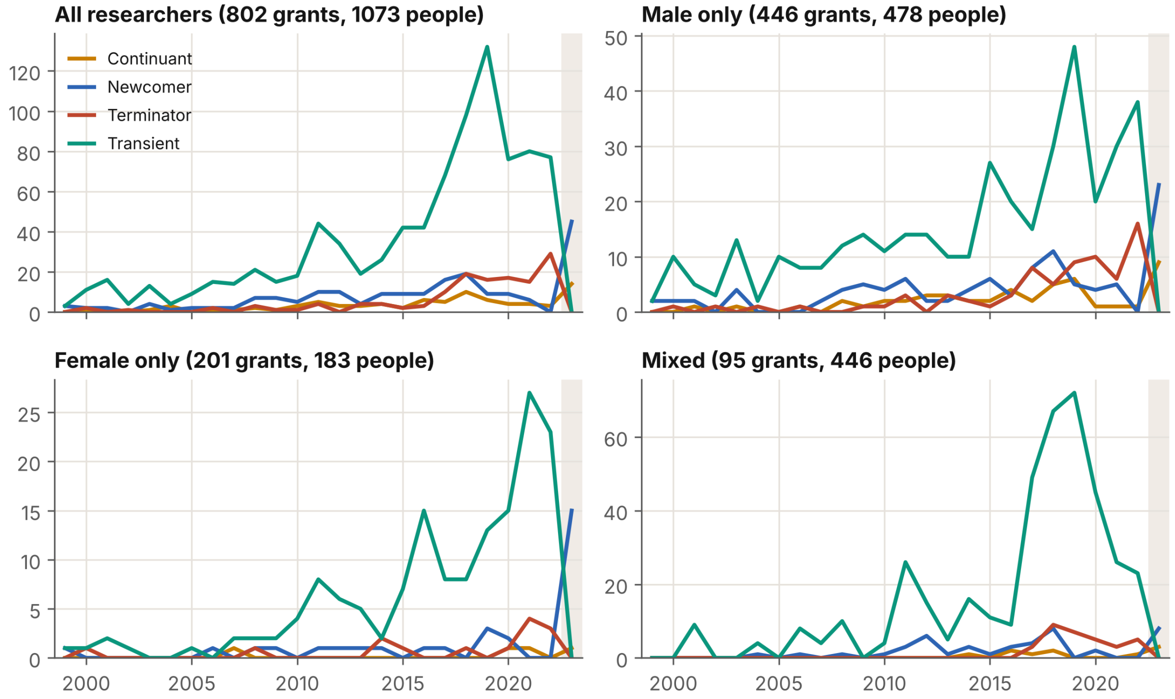


Researchers per year by engagement label, from grant start years. Panels b–d keep only grants of that team type, and labels are computed within those grants. Every researcher active in 2023 (shaded, partial year) is labelled Continuant or Newcomer because later years are not observed.

**Figure S8.** Researcher engagement over time. [FigS8_researcher_engagement]

### Figure S9. Recipient institutions per year by engagement, by topic

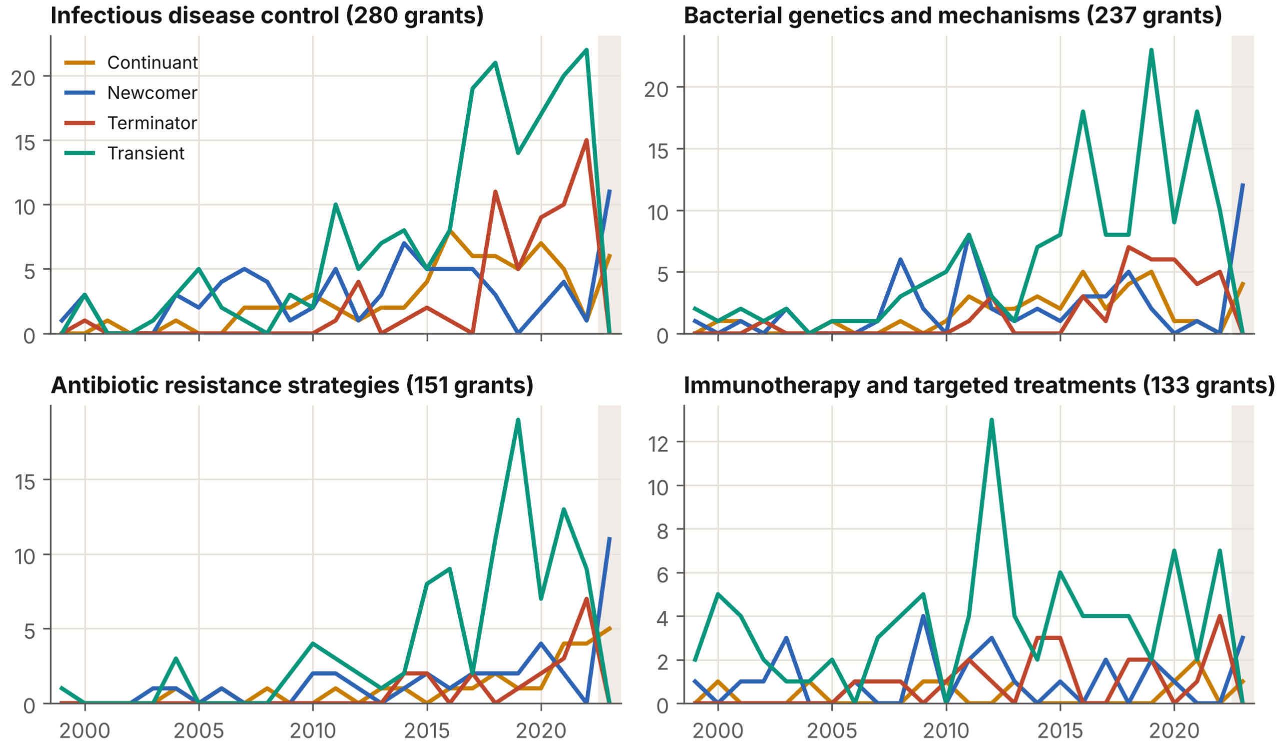


Same labels as Figure S8, applied to recipient institutions within each topic. 2023 shaded (partial year).

**Figure S9.** Institution engagement by research topic. [FigS9_institution_engagement_by_topic]

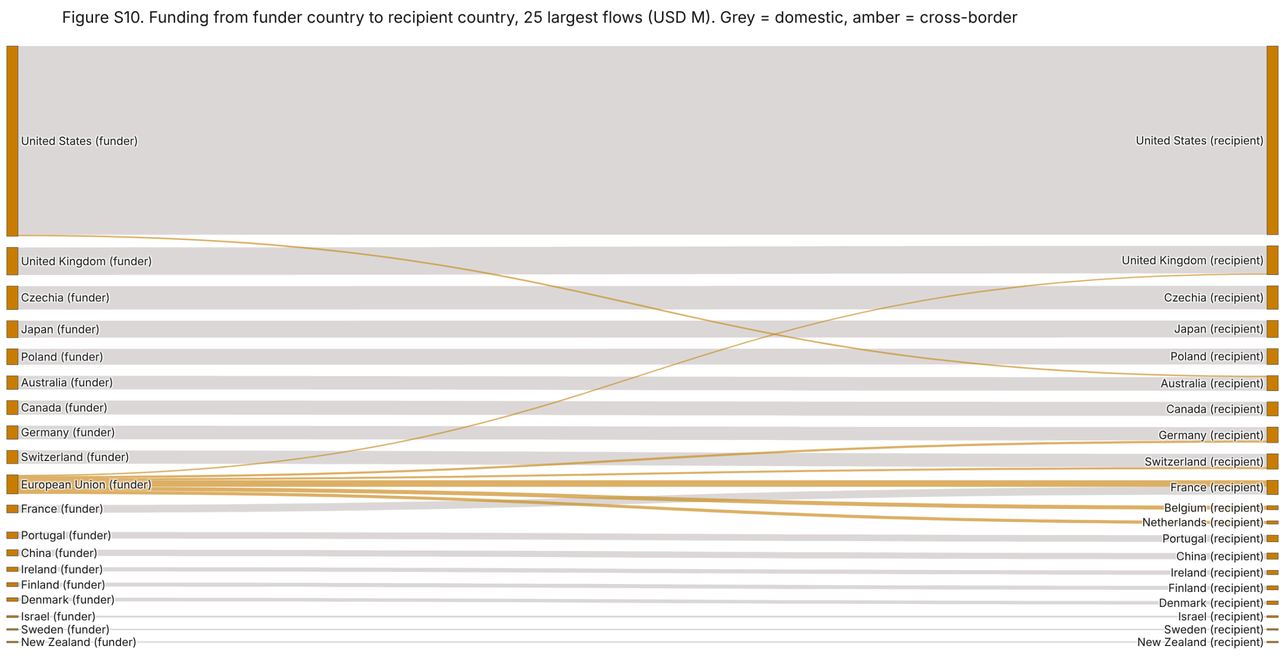


**Figure S10.** Flows of recorded funding from funder countries to recipient countries (static view of the interactive figure in the repository). [FigS10_country_flows]

**Figure S11.** Screening flow: shown as Figure 6 in the main text.

**Figure S12. Robustness of the headline results (3 of 40 checks change a conclusion; red)**

| | Funding growth (%/yr) | Global South recipients (% of funding) | Low + lower-middle income recipients (%) | US funders (% of funding) | NIAID (% of funding) | Female-only grant gap (%) | One-year-only researchers (%) | Rise in antibiotic-resistance share, 1999-2005 to 2016-2023 (points) |
|---|---|---|---|---|---|---|---|---|
| Main analysis | **16.6** | **2.1** | **0.33** | **47.7** | **21.0** | **-18.7** | **82.1** | **22.9** |
| Missing amounts: imputed, low (25th percentile) | 16.3 | 2.6 | 0.55 | 47.4 | 20.1 | | | |
| Missing amounts: imputed, central (median) | 16.0 | 3.8 | 0.54 | 45.1 | 18.9 | | | |
| Missing amounts: imputed, high (75th percentile) | 16.1 | 5.5 | 0.52 | 41.4 | 16.7 | | | |
| Count of grants instead of dollars (all 1,074) | | **17.5** | **1.4** | | | | | |
| Projects instead of records | | | | | | | 82.9 | |
| Full-span engagement rule | | | | | | | **3.5** | |
| Researchers first seen 2001-2019 | | | | | | | 83.3 | |
| Researchers who held only one grant | | | | | | | 83.9 | |
| Amounts spread over grant years | 17.1 | | | | | | | |
| Income group at grant start | | | 0.46 | | | | | |
| OECD DAC list instead of Global South | | 2.2 | | | | | | |
| Constant 2023 dollars | 14.1 | | | | | | | |
| Without NIAID | 16.2 | | | 33.8 | | | | |
| Gender: without funders in Japan and China | | | | | | -20.9 | | |
| Gender: without training awards | | | | | | -20.2 | | |
| Gender: amount per year | | | | | | -21.6 | | |
| Gender: simple model | | | | | | -27.3 | | |
| Topics: refit model, split by probability | | | | | | | | 32.5 |
| | holds if > 10 | holds if < 10 | holds if < 1 | holds if > 30, largest country | holds if > 15, largest funder | holds if < 0 | holds if > 50 | holds if > 10 |

**Figure S12.** Robustness of the headline results across 40 sensitivity checks. [FigS12_robustness]

### Supplementary Tables

**Table S1.** Completeness of the analysis table (802 grants): fields present and missing. [S1_missingness]

| | Present | Missing | Missing (%) |
|---|---|---|---|
| Grant ID | 802 | 0 | 0 |
| Title | 802 | 0 | 0 |
| Abstract | 712 | 90 | 11.2 |
| Keywords | 404 | 398 | 49.6 |
| Funding amount in USD | 802 | 0 | 0 |
| Start Year | 802 | 0 | 0 |
| End Year | 777 | 25 | 3.1 |
| Researchers | 743 | 59 | 7.4 |
| Research Organization - standardized | 802 | 0 | 0 |
| Country of standardized research organization | 802 | 0 | 0 |
| Funder | 802 | 0 | 0 |
| Funder Country | 802 | 0 | 0 |

**Table S2.** Annual growth of grants and recorded funding, 1999–2022: log-linear estimate with 95% CI, and endpoint compound annual growth rate. [S2_growth]

| | Annual growth (%) | 95% CI low | 95% CI high | Years | Endpoint CAGR 1999-2022 (%) |
|---|---|---|---|---|---|
| Grants | 12.2 | 10 | 14.5 | 1999-2022 | 10.5 |
| Funding (USD) | 16.6 | 12.6 | 20.7 | 1999-2022 | 16.3 |

**Table S3.** Grant size by team gender: Mann–Whitney tests and regression models of log(amount). The main model adjusts for team size, grant length, start year, funder and topic (HC1 robust errors). [S3_gender_tests]

| Comparison | Model | Mann-Whitney p | Adjusted difference (%) | 95% CI low (%) | 95% CI high (%) | Regression p | N |
|---|---|---|---|---|---|---|---|
| Female only vs Male only | Main: team size, grant length, start year, funder, topic | 0.047 | -18.731 | -32.289 | -2.458 | 0.026 | 742 |
| Mixed vs Male only | Main: team size, grant length, start year, funder, topic | 0.045 | -7.004 | -29.891 | 23.355 | 0.614 | 742 |
| Female only vs Male only | Amount per year of grant (team size, start year, funder, topic) | 0.047 | -21.651 | -34.814 | -5.831 | 0.009 | 742 |
| Mixed vs Male only | Amount per year of grant (team size, start year, funder, topic) | 0.045 | -5.607 | -29.117 | 25.7 | 0.693 | 742 |
| Female only vs Male only | Simple model: team size, US funder, start year | 0.047 | -27.275 | -42.739 | -7.635 | 0.009 | 742 |
| Mixed vs Male only | Simple model: team size, US funder, start year | 0.045 | 58.401 | 7.86 | 132.6 | 0.019 | 742 |

**Table S4.** Share of recorded funding by Global North and Global South, for funders and recipients (fractional counting). [S4_north_south]

| | Funders (%) | Recipients, fractional (%) |
|---|---|---|
| Global North | 98.3 | 97.9 |
| Global South | 1.7 | 2.1 |

**Table S5.** Recorded funding by country, as funder country and as recipient country (fractional counting). [S5_countries]

| | Funder (USD M) | Recipient, fractional (USD M) |
|---|---|---|
| United States | 151.8 | 151.3 |
| United Kingdom | 25.62 | 22.95 |
| Czechia | 18.58 | 18.58 |
| Japan | 13.46 | 13.46 |
| Germany | 10.65 | 12.97 |
| Switzerland | 12.27 | 12.62 |
| Poland | 12.54 | 12.54 |
| Australia | 11.01 | 12.06 |
| France | 6.29 | 11.98 |
| Canada | 11.13 | 11.68 |
| Portugal | 5.46 | 5.55 |

| | Funder (USD M) | Recipient, fractional (USD M) |
|---|---|---|
| China | 5.01 | 4.91 |
| Ireland | 3.39 | 4.19 |
| Belgium | 0 | 3.95 |
| Denmark | 2.78 | 3.5 |
| Netherlands | 0.88 | 3.37 |
| Finland | 3.16 | 3.16 |
| Israel | 1.43 | 1.6 |
| Sweden | 1.13 | 1.15 |
| New Zealand | 1.13 | 1.13 |
| Spain | 0.17 | 0.87 |
| India | 0.56 | 0.81 |
| Taiwan | 0 | 0.68 |
| Slovakia | 0.66 | 0.66 |
| Norway | 0.62 | 0.64 |
| Thailand | 0 | 0.31 |
| Italy | 0 | 0.3 |
| Georgia | 0 | 0.28 |
| Greece | 0 | 0.24 |
| Austria | 0 | 0.2 |
| Russia | 0 | 0.18 |
| South Africa | 0 | 0.17 |
| Brazil | 0 | 0.15 |
| Uganda | 0 | 0.15 |
| Singapore | 0 | 0.14 |
| Tanzania | 0 | 0.07 |
| Hungary | 0 | 0.05 |
| Ukraine | 0 | 0.04 |
| Latvia | 0.02 | 0.02 |
| European Union | 18.82 | 0 |

**Table S6.** Ten funders with the most grants. [S6_funders_by_count]

| Funder | Grants |
|---|---|
| Japan Society for the Promotion of Science | 105 |
| National Institute of Allergy and Infectious Diseases | 81 |
| National Natural Science Foundation of China | 79 |
| Natural Sciences and Engineering Research Council | 43 |
| Fundação para a Ciência e Tecnologia | 31 |
| National Science Center | 27 |
| Swiss National Science Foundation | 26 |
| National Institute of Food and Agriculture | 24 |
| Biotechnology and Biological Sciences Research Council | 24 |
| Congressionally Directed Medical Research Programs | 21 |

**Table S7.** Researcher engagement labels when a grant counts in its start year only, or in every year it runs. [S7_engagement_definitions]

| Label | Start year only (n) | Full grant span (n) | Start year only (%) | Full grant span (%) |
|---|---|---|---|---|
| Continuant | 14 | 291 | 1.3 | 27.1 |
| Newcomer | 45 | 45 | 4.2 | 4.2 |
| Terminator | 133 | 699 | 12.4 | 65.1 |
| Transient | 881 | 38 | 82.1 | 3.5 |

**Table S8.** Mean c_v topic coherence (top 30 words) for LDA models with k = 2–12 topics. [S8_topic_coherence]

| k | c_v coherence |
|---|---|
| 2 | 0.347 |
| 3 | 0.369 |
| 4 | 0.366 |

| k | c_v coherence |
|---|---|
| 5 | 0.373 |
| 6 | 0.353 |
| 7 | 0.359 |
| 8 | 0.344 |
| 9 | 0.339 |
| 10 | 0.322 |
| 11 | 0.316 |
| 12 | 0.335 |

**Table S9.** Topic stability: share of grants that keep the same topic when the model is refitted with other random seeds. [S9_topic_stability]

| k | Seed | Same topic (%) |
|---|---|---|
| 4 | 1 | 56.7 |
| 4 | 2 | 67.5 |
| 4 | 3 | 63.6 |
| 4 | 4 | 52.2 |
| 4 | 5 | 61.8 |
| 5 | 1 | 59.6 |
| 5 | 2 | 54.9 |
| 5 | 3 | 68.1 |
| 5 | 4 | 64.7 |
| 5 | 5 | 51.2 |
| 6 | 1 | 57.9 |
| 6 | 2 | 62.2 |
| 6 | 3 | 59.8 |
| 6 | 4 | 63.3 |
| 6 | 5 | 53.4 |

**Table S10.** Top 50 words of each topic in the refitted k = 5 model (notebook 02). Refitted topics are matched to the topics used in the paper by the best one-to-one overlap of grant assignments (59.1% of grants agree); the fifth refitted topic corresponds to the paper's single unclassified grant only loosely. [S10_topic_top_words]

| Rank | Bacterial genetics and mechanisms | Antibiotic resistance strategies | Immunotherapy and targeted treatments | Infectious disease control | Unclassified |
|---|---|---|---|---|---|
| 1 | gene | infection | peptide | bacteriophage | membrane |
| 2 | cell | antibiotic | cell | bacteria | antibacterial |
| 3 | protein | bacteria | cancer | treatment | bacteria |
| 4 | bacteriophage | resistance | antibody | infection | negative |
| 5 | host | bacterial | display | therapy | enzyme |
| 6 | bacterial | bacteriophage | target | bacterial | positive |
| 7 | study | therapy | protein | strain | agent |
| 8 | target | resistant | vaccine | resistant | resistant |
| 9 | research | pathogen | library | antibiotic | bacterial |
| 10 | system | system | bind | control | gene |
| 11 | therapy | therapeutic | specific | food | resistance |
| 12 | enzyme | model | antibodie | pathogen | release |
| 13 | activity | research | disease | develop | analyze |
| 14 | project | human | human | disease | activity |
| 15 | specific | clinical | therapeutic | biofilm | expect |
| 16 | virus | study | response | project | bacterium |
| 17 | disease | treatment | treatment | isolate | contain |
| 18 | genetic | host | develop | study | strain |
| 19 | mechanism | disease | antigen | antimicrobial | induce |
| 20 | expression | development | identify | method | property |
| 21 | bacteria | health | therapy | infect | analysis |
| 22 | genome | develop | patient | alternative | derive |
| 23 | method | antimicrobial | receptor | lytic | method |
| 24 | development | project | anti | development | genome |

| Rank | Bacterial genetics and mechanisms | Antibiotic resistance strategies | Immunotherapy and targeted treatments | Infectious disease control | Unclassified |
|---|---|---|---|---|---|
| 25 | process | virus | screen | product | obtain |
| 26 | lytic | mechanism | study | animal | information |
| 27 | delivery | patient | development | result | production |
| 28 | develop | cell | system | reduce | form |
| 29 | molecular | interaction | immune | resistance | select |
| 30 | technology | immune | tissue | treat | isolate |
| 31 | viral | strain | high | specific | carry |
| 32 | base | work | technology | target | possible |
| 33 | dna | infect | base | base | generation |
| 34 | infection | kill | molecule | application | increase |
| 35 | sequence | pseudomona | method | increase | spread |
| 36 | result | aim | research | cocktail | environment |
| 37 | analysis | treat | mouse | wound | produce |
| 38 | application | base | vivo | effect | study |
| 39 | structure | understand | clinical | effective | virus |
| 40 | treatment | target | aim | potential | promote |
| 41 | therapeutic | approach | effect | environment | identify |
| 42 | production | aeruginosa | molecular | host | year |
| 43 | interaction | agent | type | test | mechanism |
| 44 | receptor | provide | application | production | emergence |
| 45 | bind | goal | model | virus | surface |
| 46 | provide | biofilm | approach | research | water |
| 47 | surface | datum | provide | technology | perform |
| 48 | platform | animal | surface | safety | consider |
| 49 | high | genetic | project | efficacy | report |
| 50 | control | gene | efficacy | staphylococcus | research |

**Table S11.** Global South recipient countries: recipient entries and recorded funding (fractional counting). [S11_global_south_recipients]

| Country | Recipient entries | USD (M) | Share of all funding (%) |
|---|---|---|---|
| China | 84 | 4.91 | 1.54 |
| India | 13 | 0.81 | 0.25 |
| Thailand | 3 | 0.31 | 0.1 |
| South Africa | 2 | 0.17 | 0.05 |
| Brazil | 1 | 0.15 | 0.05 |
| Uganda | 2 | 0.15 | 0.05 |
| Singapore | 2 | 0.14 | 0.04 |
| Tanzania | 1 | 0.07 | 0.02 |
| Total Global South | 108 | 6.72 | 2.11 |

**Table S12.** Domestic and cross-border funding. A grant is domestic when every recipient institution is in the funder's country. [S12_domestic_vs_crossborder]

| Funding type | Grants | USD (M) | Share of funding (%) |
|---|---|---|---|
| Cross-border | 65 | 36.1 | 11.3 |
| Domestic | 737 | 282.5 | 88.7 |

**Table S13.** Largest cross-border funders. [S13_crossborder_funders]

| Funder | Cross-border grants | USD (M) |
|---|---|---|
| European Commission | 19 | 14.64 |
| Biotechnology and Biological Sciences Research Council | 12 | 5.6 |
| Swiss National Science Foundation | 8 | 5.12 |
| European Research Council | 4 | 4.18 |
| Engineering and Physical Sciences Research Council | 1 | 2.02 |
| National Institute of Allergy and Infectious Diseases | 5 | 1.91 |
| Australian Research Council | 2 | 0.83 |
| Medical Research Council | 1 | 0.61 |

**Table S14.** Grants per start year for the 15 funders with the most grants (data for Figure S2). [S14_funder_by_year]

| Funder | 1999 | 2000 | 2001 | 2002 | 2003 | 2004 | 2005 | 2006 | 2007 | 2008 | 2009 | 2010 | 2011 | 2012 | 2013 | 2014 | 2015 | 2016 | 2017 | 2018 | 2019 | 2020 | 2021 | 2022 | 2023 |
|---|---|---|---|---|---|---|---|---|---|---|---|---|---|---|---|---|---|---|---|---|---|---|---|---|---|
| Japan Society for the Promotion of Science | 0 | 2 | 2 | 0 | 5 | 2 | 2 | 3 | 1 | 4 | 0 | 2 | 5 | 1 | 1 | 3 | 3 | 3 | 5 | 7 | 13 | 9 | 6 | 13 | 13 |
| National Institute of Allergy and Infectious Diseases | 0 | 1 | 0 | 1 | 0 | 0 | 0 | 1 | 2 | 1 | 2 | 1 | 0 | 1 | 0 | 0 | 10 | 2 | 3 | 7 | 4 | 11 | 17 | 9 | 8 |
| National Natural Science Foundation of China | 0 | 1 | 1 | 2 | 0 | 0 | 1 | 0 | 0 | 4 | 4 | 1 | 2 | 7 | 3 | 4 | 8 | 6 | 8 | 5 | 13 | 8 | 1 | 0 | 0 |
| Natural Sciences and Engineering Research Council | 2 | 3 | 1 | 1 | 0 | 0 | 0 | 1 | 1 | 1 | 1 | 0 | 1 | 2 | 0 | 1 | 3 | 2 | 2 | 6 | 2 | 3 | 8 | 2 | 0 |
| Fundação para a Ciência e Tecnologia | 0 | 0 | 0 | 0 | 0 | 0 | 0 | 0 | 2 | 0 | 0 | 3 | 1 | 2 | 4 | 2 | 2 | 1 | 1 | 3 | 1 | 2 | 3 | 3 | 1 |
| National Science Center | 0 | 0 | 0 | 0 | 0 | 0 | 0 | 0 | 0 | 0 | 0 | 0 | 0 | 1 | 3 | 1 | 1 | 1 | 3 | 4 | 6 | 2 | 3 | 2 | 0 |
| Swiss National Science Foundation | 0 | 0 | 1 | 0 | 0 | 0 | 0 | 1 | 0 | 0 | 0 | 1 | 0 | 0 | 0 | 4 | 0 | 2 | 2 | 3 | 2 | 3 | 1 | 3 | 3 |
| National Institute of Food and Agriculture | 0 | 1 | 1 | 0 | 0 | 0 | 0 | 0 | 1 | 0 | 0 | 0 | 0 | 1 | 0 | 0 | 2 | 1 | 1 | 2 | 4 | 0 | 7 | 1 | 2 |
| Biotechnology and Biological Sciences Research Council | 0 | 0 | 0 | 0 | 0 | 0 | 0 | 1 | 0 | 0 | 0 | 1 | 3 | 2 | 1 | 0 | 2 | 1 | 2 | 2 | 1 | 4 | 2 | 0 | 2 |
| Congressionally Directed Medical Research Programs | 2 | 2 | 2 | 0 | 0 | 0 | 0 | 1 | 0 | 0 | 3 | 0 | 0 | 2 | 0 | 0 | 0 | 3 | 2 | 3 | 0 | 1 | 0 | 0 | 0 |
| European Commission | 1 | 0 | 0 | 0 | 0 | 2 | 1 | 0 | 0 | 0 | 0 | 0 | 1 | 1 | 1 | 0 | 0 | 1 | 0 | 2 | 3 | 1 | 1 | 1 | 3 |
| Canadian Institutes of Health Research | 0 | 0 | 0 | 0 | 0 | 0 | 1 | 0 | 1 | 3 | 0 | 1 | 0 | 1 | 0 | 0 | 0 | 0 | 2 | 1 | 0 | 1 | 1 | 2 | 1 |
| Directorate for Technology, Innovation and Partnerships | 0 | 0 | 0 | 0 | 0 | 0 | 0 | 0 | 0 | 0 | 0 | 2 | 1 | 0 | 0 | 0 | 0 | 0 | 2 | 1 | 1 | 1 | 4 | 1 | 0 |
| National Cancer Institute | 0 | 2 | 0 | 0 | 0 | 1 | 0 | 0 | 1 | 0 | 2 | 0 | 1 | 1 | 0 | 0 | 0 | 1 | 0 | 1 | 0 | 1 | 0 | 1 | 0 |
| National Institute of General Medical Sciences | 0 | 0 | 1 | 0 | 0 | 0 | 0 | 0 | 0 | 0 | 1 | 0 | 0 | 1 | 0 | 0 | 0 | 1 | 0 | 0 | 2 | 0 | 2 | 2 | 1 |

**Table S15.** Topic shares (% of each funder's grants) for the 15 funders with the most grants (data for Figure S3). [S15_topic_share_by_funder]

| Funder | Infectious disease control | Bacterial genetics and mechanisms | Antibiotic resistance strategies | Immunotherapy and targeted treatments |
|---|---|---|---|---|
| Japan Society for the Promotion of Science | 32.4 | 27.6 | 24.8 | 15.2 |
| National Institute of Allergy and Infectious Diseases | 7.4 | 29.6 | 58 | 4.9 |
| National Natural Science Foundation of China | 5.1 | 38.5 | 12.8 | 43.6 |
| Natural Sciences and Engineering Research Council | 41.9 | 44.2 | 4.7 | 9.3 |
| Fundação para a Ciência e Tecnologia | 67.7 | 9.7 | 12.9 | 9.7 |
| National Science Center | 22.2 | 70.4 | 3.7 | 3.7 |
| Swiss National Science Foundation | 34.6 | 34.6 | 26.9 | 3.8 |
| National Institute of Food and Agriculture | 75 | 20.8 | 4.2 | 0 |
| Biotechnology and Biological Sciences Research Council | 29.2 | 50 | 12.5 | 8.3 |
| Congressionally Directed Medical Research Programs | 33.3 | 0 | 4.8 | 61.9 |
| European Commission | 42.1 | 36.8 | 15.8 | 5.3 |
| Canadian Institutes of Health Research | 53.3 | 40 | 0 | 6.7 |
| Directorate for Technology, Innovation and Partnerships | 61.5 | 15.4 | 7.7 | 15.4 |
| National Cancer Institute | 0 | 8.3 | 8.3 | 83.3 |
| National Institute of General Medical Sciences | 0 | 72.7 | 9.1 | 18.2 |

**Table S16.** Topic shares (% of each country's grants) for the 15 largest recipient countries (data for Figure S4). [S16_topic_share_by_recipient]

| | Infectious disease control | Bacterial genetics and mechanisms | Antibiotic resistance strategies | Immunotherapy and targeted treatments |
|---|---|---|---|---|
| United States | 25.4 | 26.1 | 26.6 | 21.8 |
| Japan | 33.3 | 27 | 24.3 | 15.3 |
| China | 6.3 | 38.6 | 13.3 | 41.8 |
| Canada | 47 | 39.6 | 5.4 | 8.1 |
| United Kingdom | 39.1 | 39.1 | 10.9 | 10.9 |
| Poland | 28.6 | 57.1 | 8.6 | 5.7 |
| Portugal | 71.2 | 9.4 | 12.5 | 7 |
| Switzerland | 35.4 | 32.1 | 24.1 | 8.5 |
| Australia | 84.4 | 1 | 4.9 | 9.8 |
| Czechia | 41.2 | 23.5 | 35.3 | 0 |
| Germany | 60.5 | 20.4 | 19.1 | 0 |
| France | 44.2 | 28 | 15.3 | 12.4 |

| | Infectious disease control | Bacterial genetics and mechanisms | Antibiotic resistance strategies | Immunotherapy and targeted treatments |
|---|---|---|---|---|
| Global North | 37.8 | 29.1 | 19.3 | 13.8 |
| Global South | 13.6 | 33 | 15.5 | 37.8 |
| All grants | 35 | 29.6 | 18.9 | 16.6 |

**Table S17.** Most distinctive words of each topic (data for Figure S5). [S17_topic_distinctive_words]

| Infectious disease control | Bacterial genetics and mechanisms | Antibiotic resistance strategies | Immunotherapy and targeted treatments |
|---|---|---|---|
| infections | phages | resistance | cancer |
| bacteriophage | host | biofilm | peptide |
| phages | resistance | clinical | display |
| resistant | bacteriophage | infections | antibody |
| treatment | protein | infection | antibodies |
| antibiotic | genes | phages | tumor |
| antibiotics | antibiotic | antibiotic | peptides |
| project | gene | model | cells |
| food | infection | resistant | gene |
| resistance | proteins | aureus | protein |
| antimicrobial | cell | therapeutic | cell |
| development | dna | lytic | vaccine |
| alternative | research | development | specific |
| strains | resistant | biofilms | library |
| coli | interactions | treatment | breast |
| based | systems | antibiotics | targeting |
| infection | project | bacteriophage | delivery |
| fish | mechanisms | disease | receptor |
| technology | lytic | staphylococcus | targeted |
| pathogens | genetic | pathogen | therapeutic |
| health | crispr | models | technology |
| research | antibiotics | animal | brain |
| control | biology | aeruginosa | binding |
| poultry | molecular | efficacy | proteins |
| production | antibacterial | pneumoniae | libraries |
| disease | coli | testing | human |
| develop | genome | vivo | screening |
| viruses | study | baumannii | prostate |
| treat | mechanism | mdr | research |
| products | activity | pseudomonas | disease |

**Table S18.** Jaccard similarity between the top-word sets of the topics (data for Figure S5). [S18_topic_jaccard]

| | Infectious disease control | Bacterial genetics and mechanisms | Antibiotic resistance strategies | Immunotherapy and targeted treatments |
|---|---|---|---|---|
| Infectious disease control | 1 | 0.2 | 0.224 | 0.053 |
| Bacterial genetics and mechanisms | 0.2 | 1 | 0.154 | 0.091 |
| Antibiotic resistance strategies | 0.224 | 0.154 | 1 | 0.034 |
| Immunotherapy and targeted treatments | 0.053 | 0.091 | 0.034 | 1 |

**Table S19.** Named researchers by inferred gender and start year (data for Figure S6). [S19_researchers_by_gender_year]

| Year | Male | Female | Unknown | Female share (%) |
|---|---|---|---|---|
| 1,999 | 4 | 2 | 0 | 33.3 |
| 2,000 | 13 | 2 | 0 | 13.3 |
| 2,001 | 15 | 4 | 0 | 21.1 |
| 2,002 | 4 | 1 | 0 | 20 |
| 2,003 | 18 | 0 | 0 | 0 |
| 2,004 | 7 | 1 | 0 | 12.5 |
| 2,005 | 10 | 1 | 0 | 9.1 |
| 2,006 | 15 | 5 | 0 | 25 |

| Year | Male | Female | Unknown | Female share (%) |
|---|---|---|---|---|
| 2,007 | 12 | 5 | 0 | 29.4 |
| 2,008 | 27 | 6 | 0 | 18.2 |
| 2,009 | 21 | 3 | 0 | 12.5 |
| 2,010 | 20 | 7 | 0 | 25.9 |
| 2,011 | 36 | 27 | 0 | 42.9 |
| 2,012 | 30 | 17 | 0 | 36.2 |
| 2,013 | 20 | 9 | 1 | 31 |
| 2,014 | 29 | 13 | 1 | 31 |
| 2,015 | 44 | 11 | 0 | 20 |
| 2,016 | 40 | 20 | 0 | 33.3 |
| 2,017 | 65 | 34 | 0 | 34.3 |
| 2,018 | 92 | 54 | 0 | 37 |
| 2,019 | 109 | 54 | 0 | 33.1 |
| 2,020 | 65 | 41 | 0 | 38.7 |
| 2,021 | 59 | 44 | 2 | 42.7 |
| 2,022 | 70 | 39 | 0 | 35.8 |
| 2,023 | 39 | 20 | 0 | 33.9 |

**Table S20.** Team gender composition by topic, % of grants (data for Figure S7). [S20_team_gender_by_topic]

| Topic | Male only | Mixed | Female only | Gender unknown |
|---|---|---|---|---|
| Infectious disease control | 53.6 | 11.8 | 24.3 | 10.4 |
| Bacterial genetics and mechanisms | 51.5 | 13.1 | 29.5 | 5.9 |
| Antibiotic resistance strategies | 56.3 | 15.2 | 19.9 | 8.6 |
| Immunotherapy and targeted treatments | 66.2 | 6 | 24.8 | 3 |
| All grants | 55.6 | 11.8 | 25.1 | 7.5 |

**Table S21.** Researcher engagement labels by year (data for Figure S8). [S21_researcher_engagement_by_year]

| Group | Year | Continuant | Newcomer | Terminator | Transient |
|---|---|---|---|---|---|
| All researchers | 1999 | 0 | 3 | 0 | 3 |
| All researchers | 2000 | 0 | 2 | 2 | 11 |
| All researchers | 2001 | 1 | 2 | 0 | 16 |
| All researchers | 2002 | 0 | 0 | 1 | 4 |
| All researchers | 2003 | 1 | 4 | 0 | 13 |
| All researchers | 2004 | 3 | 1 | 0 | 4 |
| All researchers | 2005 | 0 | 2 | 0 | 9 |
| All researchers | 2006 | 1 | 2 | 2 | 15 |
| All researchers | 2007 | 1 | 2 | 0 | 14 |
| All researchers | 2008 | 2 | 7 | 3 | 21 |
| All researchers | 2009 | 1 | 7 | 1 | 15 |
| All researchers | 2010 | 3 | 5 | 1 | 18 |
| All researchers | 2011 | 5 | 10 | 4 | 44 |
| All researchers | 2012 | 3 | 10 | 0 | 34 |
| All researchers | 2013 | 3 | 4 | 4 | 19 |
| All researchers | 2014 | 4 | 9 | 4 | 26 |
| All researchers | 2015 | 2 | 9 | 2 | 42 |
| All researchers | 2016 | 6 | 9 | 3 | 42 |
| All researchers | 2017 | 5 | 16 | 10 | 68 |
| All researchers | 2018 | 10 | 19 | 19 | 98 |
| All researchers | 2019 | 6 | 9 | 16 | 132 |
| All researchers | 2020 | 4 | 9 | 17 | 76 |
| All researchers | 2021 | 4 | 6 | 15 | 80 |
| All researchers | 2022 | 3 | 0 | 29 | 77 |
| All researchers | 2023 | 14 | 45 | 0 | 0 |
| Male only | 1999 | 0 | 2 | 0 | 2 |
| Male only | 2000 | 0 | 2 | 1 | 10 |

| Group | Year | Continuant | Newcomer | Terminator | Transient |
|---|---|---|---|---|---|
| Male only | 2001 | 1 | 2 | 0 | 5 |
| Male only | 2002 | 0 | 0 | 1 | 3 |
| Male only | 2003 | 1 | 4 | 0 | 13 |
| Male only | 2004 | 0 | 0 | 1 | 2 |
| Male only | 2005 | 0 | 0 | 0 | 10 |
| Male only | 2006 | 1 | 0 | 1 | 8 |
| Male only | 2007 | 0 | 2 | 0 | 8 |
| Male only | 2008 | 2 | 4 | 0 | 12 |
| Male only | 2009 | 1 | 5 | 1 | 14 |
| Male only | 2010 | 2 | 4 | 1 | 11 |
| Male only | 2011 | 2 | 6 | 3 | 14 |
| Male only | 2012 | 3 | 2 | 0 | 14 |
| Male only | 2013 | 3 | 2 | 3 | 10 |
| Male only | 2014 | 2 | 4 | 2 | 10 |
| Male only | 2015 | 2 | 6 | 1 | 27 |
| Male only | 2016 | 4 | 3 | 3 | 20 |
| Male only | 2017 | 2 | 8 | 8 | 15 |
| Male only | 2018 | 5 | 11 | 5 | 30 |
| Male only | 2019 | 6 | 5 | 9 | 48 |
| Male only | 2020 | 1 | 4 | 10 | 20 |
| Male only | 2021 | 1 | 5 | 6 | 30 |
| Male only | 2022 | 1 | 0 | 16 | 38 |
| Male only | 2023 | 9 | 23 | 0 | 0 |
| Female only | 1999 | 0 | 1 | 0 | 1 |
| Female only | 2000 | 0 | 0 | 1 | 1 |
| Female only | 2001 | 0 | 0 | 0 | 2 |
| Female only | 2002 | 0 | 0 | 0 | 1 |
| Female only | 2003 | 0 | 0 | 0 | 0 |
| Female only | 2004 | 0 | 0 | 0 | 0 |
| Female only | 2005 | 0 | 0 | 0 | 1 |
| Female only | 2006 | 0 | 1 | 0 | 0 |
| Female only | 2007 | 1 | 0 | 0 | 2 |
| Female only | 2008 | 0 | 1 | 1 | 2 |
| Female only | 2009 | 0 | 1 | 0 | 2 |
| Female only | 2010 | 0 | 0 | 0 | 4 |
| Female only | 2011 | 0 | 1 | 0 | 8 |
| Female only | 2012 | 0 | 1 | 0 | 6 |
| Female only | 2013 | 0 | 1 | 0 | 5 |
| Female only | 2014 | 0 | 1 | 2 | 2 |
| Female only | 2015 | 0 | 0 | 1 | 7 |
| Female only | 2016 | 0 | 1 | 0 | 15 |
| Female only | 2017 | 0 | 1 | 0 | 8 |
| Female only | 2018 | 0 | 0 | 1 | 8 |
| Female only | 2019 | 0 | 3 | 0 | 13 |
| Female only | 2020 | 1 | 2 | 1 | 15 |
| Female only | 2021 | 1 | 0 | 4 | 27 |
| Female only | 2022 | 0 | 0 | 3 | 23 |
| Female only | 2023 | 1 | 15 | 0 | 0 |
| Mixed | 1999 | 0 | 0 | 0 | 0 |
| Mixed | 2000 | 0 | 0 | 0 | 0 |
| Mixed | 2001 | 0 | 0 | 0 | 9 |
| Mixed | 2002 | 0 | 0 | 0 | 0 |
| Mixed | 2003 | 0 | 0 | 0 | 0 |
| Mixed | 2004 | 0 | 1 | 0 | 4 |
| Mixed | 2005 | 0 | 0 | 0 | 0 |
| Mixed | 2006 | 0 | 1 | 0 | 8 |

| Group | Year | Continuant | Newcomer | Terminator | Transient |
|---|---|---|---|---|---|
| Mixed | 2007 | 0 | 0 | 0 | 4 |
| Mixed | 2008 | 0 | 1 | 0 | 10 |
| Mixed | 2009 | 0 | 0 | 0 | 0 |
| Mixed | 2010 | 0 | 1 | 0 | 4 |
| Mixed | 2011 | 0 | 3 | 0 | 26 |
| Mixed | 2012 | 0 | 6 | 0 | 15 |
| Mixed | 2013 | 0 | 1 | 0 | 5 |
| Mixed | 2014 | 1 | 3 | 0 | 16 |
| Mixed | 2015 | 0 | 1 | 0 | 11 |
| Mixed | 2016 | 2 | 3 | 0 | 9 |
| Mixed | 2017 | 1 | 4 | 3 | 49 |
| Mixed | 2018 | 2 | 8 | 9 | 67 |
| Mixed | 2019 | 0 | 0 | 7 | 72 |
| Mixed | 2020 | 0 | 2 | 5 | 45 |
| Mixed | 2021 | 0 | 0 | 3 | 26 |
| Mixed | 2022 | 1 | 0 | 5 | 23 |
| Mixed | 2023 | 3 | 8 | 0 | 0 |

**Table S22.** Institution engagement labels by topic and year (data for Figure S9). [S22_institution_engagement_by_topic]

| Topic | Year | Continuant | Newcomer | Terminator | Transient |
|---|---|---|---|---|---|
| Infectious disease control | 1999 | 0 | 1 | 0 | 0 |
| Infectious disease control | 2000 | 0 | 3 | 1 | 3 |
| Infectious disease control | 2001 | 1 | 0 | 0 | 0 |
| Infectious disease control | 2002 | 0 | 0 | 0 | 0 |
| Infectious disease control | 2003 | 0 | 0 | 0 | 1 |
| Infectious disease control | 2004 | 1 | 3 | 0 | 3 |
| Infectious disease control | 2005 | 0 | 2 | 0 | 5 |
| Infectious disease control | 2006 | 0 | 4 | 0 | 2 |
| Infectious disease control | 2007 | 2 | 5 | 0 | 1 |
| Infectious disease control | 2008 | 2 | 4 | 0 | 0 |
| Infectious disease control | 2009 | 2 | 1 | 0 | 3 |
| Infectious disease control | 2010 | 3 | 2 | 0 | 2 |
| Infectious disease control | 2011 | 2 | 5 | 1 | 10 |
| Infectious disease control | 2012 | 1 | 1 | 4 | 5 |
| Infectious disease control | 2013 | 2 | 3 | 0 | 7 |
| Infectious disease control | 2014 | 2 | 7 | 1 | 8 |
| Infectious disease control | 2015 | 4 | 5 | 2 | 5 |
| Infectious disease control | 2016 | 8 | 5 | 1 | 8 |
| Infectious disease control | 2017 | 6 | 5 | 0 | 19 |
| Infectious disease control | 2018 | 6 | 3 | 11 | 21 |
| Infectious disease control | 2019 | 5 | 0 | 5 | 14 |
| Infectious disease control | 2020 | 7 | 2 | 9 | 17 |
| Infectious disease control | 2021 | 5 | 4 | 10 | 20 |
| Infectious disease control | 2022 | 1 | 1 | 15 | 22 |
| Infectious disease control | 2023 | 6 | 11 | 0 | 0 |
| Bacterial genetics and mechanisms | 1999 | 0 | 1 | 0 | 2 |
| Bacterial genetics and mechanisms | 2000 | 1 | 0 | 0 | 1 |
| Bacterial genetics and mechanisms | 2001 | 1 | 1 | 0 | 2 |
| Bacterial genetics and mechanisms | 2002 | 0 | 0 | 1 | 1 |
| Bacterial genetics and mechanisms | 2003 | 0 | 2 | 0 | 2 |
| Bacterial genetics and mechanisms | 2004 | 0 | 0 | 0 | 0 |
| Bacterial genetics and mechanisms | 2005 | 1 | 0 | 0 | 1 |
| Bacterial genetics and mechanisms | 2006 | 0 | 0 | 0 | 1 |
| Bacterial genetics and mechanisms | 2007 | 0 | 1 | 0 | 1 |
| Bacterial genetics and mechanisms | 2008 | 1 | 6 | 0 | 3 |
| Bacterial genetics and mechanisms | 2009 | 0 | 2 | 0 | 4 |

| Topic | Year | Continuant | Newcomer | Terminator | Transient |
|---|---|---|---|---|---|
| Bacterial genetics and mechanisms | 2010 | 1 | 0 | 0 | 5 |
| Bacterial genetics and mechanisms | 2011 | 3 | 8 | 1 | 8 |
| Bacterial genetics and mechanisms | 2012 | 2 | 2 | 3 | 3 |
| Bacterial genetics and mechanisms | 2013 | 2 | 1 | 0 | 1 |
| Bacterial genetics and mechanisms | 2014 | 3 | 2 | 0 | 7 |
| Bacterial genetics and mechanisms | 2015 | 2 | 1 | 0 | 8 |
| Bacterial genetics and mechanisms | 2016 | 5 | 3 | 3 | 18 |
| Bacterial genetics and mechanisms | 2017 | 2 | 3 | 1 | 8 |
| Bacterial genetics and mechanisms | 2018 | 4 | 5 | 7 | 8 |
| Bacterial genetics and mechanisms | 2019 | 5 | 2 | 6 | 23 |
| Bacterial genetics and mechanisms | 2020 | 1 | 0 | 6 | 9 |
| Bacterial genetics and mechanisms | 2021 | 1 | 1 | 4 | 18 |
| Bacterial genetics and mechanisms | 2022 | 0 | 0 | 5 | 10 |
| Bacterial genetics and mechanisms | 2023 | 4 | 12 | 0 | 0 |
| Antibiotic resistance strategies | 1999 | 0 | 0 | 0 | 1 |
| Antibiotic resistance strategies | 2000 | 0 | 0 | 0 | 0 |
| Antibiotic resistance strategies | 2001 | 0 | 0 | 0 | 0 |
| Antibiotic resistance strategies | 2002 | 0 | 0 | 0 | 0 |
| Antibiotic resistance strategies | 2003 | 0 | 1 | 0 | 0 |
| Antibiotic resistance strategies | 2004 | 1 | 1 | 0 | 3 |
| Antibiotic resistance strategies | 2005 | 0 | 0 | 0 | 0 |
| Antibiotic resistance strategies | 2006 | 1 | 1 | 0 | 0 |
| Antibiotic resistance strategies | 2007 | 0 | 0 | 0 | 0 |
| Antibiotic resistance strategies | 2008 | 1 | 0 | 0 | 0 |
| Antibiotic resistance strategies | 2009 | 0 | 0 | 0 | 2 |
| Antibiotic resistance strategies | 2010 | 0 | 2 | 0 | 4 |
| Antibiotic resistance strategies | 2011 | 1 | 2 | 0 | 3 |
| Antibiotic resistance strategies | 2012 | 0 | 1 | 0 | 2 |
| Antibiotic resistance strategies | 2013 | 1 | 0 | 0 | 1 |
| Antibiotic resistance strategies | 2014 | 1 | 1 | 2 | 2 |
| Antibiotic resistance strategies | 2015 | 0 | 2 | 2 | 8 |
| Antibiotic resistance strategies | 2016 | 1 | 1 | 0 | 9 |
| Antibiotic resistance strategies | 2017 | 1 | 2 | 2 | 2 |
| Antibiotic resistance strategies | 2018 | 2 | 2 | 0 | 11 |
| Antibiotic resistance strategies | 2019 | 1 | 2 | 1 | 19 |
| Antibiotic resistance strategies | 2020 | 1 | 4 | 2 | 7 |
| Antibiotic resistance strategies | 2021 | 4 | 2 | 3 | 13 |
| Antibiotic resistance strategies | 2022 | 4 | 0 | 7 | 9 |
| Antibiotic resistance strategies | 2023 | 5 | 11 | 0 | 0 |
| Immunotherapy and targeted treatments | 1999 | 0 | 1 | 0 | 2 |
| Immunotherapy and targeted treatments | 2000 | 1 | 0 | 0 | 5 |
| Immunotherapy and targeted treatments | 2001 | 0 | 1 | 0 | 4 |
| Immunotherapy and targeted treatments | 2002 | 0 | 1 | 0 | 2 |
| Immunotherapy and targeted treatments | 2003 | 0 | 3 | 0 | 1 |
| Immunotherapy and targeted treatments | 2004 | 1 | 0 | 0 | 1 |
| Immunotherapy and targeted treatments | 2005 | 0 | 0 | 0 | 2 |
| Immunotherapy and targeted treatments | 2006 | 0 | 1 | 1 | 0 |
| Immunotherapy and targeted treatments | 2007 | 0 | 0 | 1 | 3 |
| Immunotherapy and targeted treatments | 2008 | 0 | 0 | 1 | 4 |
| Immunotherapy and targeted treatments | 2009 | 1 | 4 | 0 | 5 |
| Immunotherapy and targeted treatments | 2010 | 1 | 0 | 1 | 0 |
| Immunotherapy and targeted treatments | 2011 | 0 | 2 | 2 | 4 |
| Immunotherapy and targeted treatments | 2012 | 0 | 3 | 1 | 13 |
| Immunotherapy and targeted treatments | 2013 | 1 | 1 | 0 | 4 |
| Immunotherapy and targeted treatments | 2014 | 0 | 0 | 3 | 2 |
| Immunotherapy and targeted treatments | 2015 | 0 | 1 | 3 | 6 |

| Topic | Year | Continuant | Newcomer | Terminator | Transient |
|---|---|---|---|---|---|
| Immunotherapy and targeted treatments | 2016 | 0 | 0 | 0 | 4 |
| Immunotherapy and targeted treatments | 2017 | 0 | 2 | 0 | 4 |
| Immunotherapy and targeted treatments | 2018 | 0 | 0 | 2 | 4 |
| Immunotherapy and targeted treatments | 2019 | 0 | 2 | 2 | 2 |
| Immunotherapy and targeted treatments | 2020 | 1 | 1 | 0 | 7 |
| Immunotherapy and targeted treatments | 2021 | 2 | 0 | 1 | 2 |
| Immunotherapy and targeted treatments | 2022 | 0 | 0 | 4 | 7 |
| Immunotherapy and targeted treatments | 2023 | 1 | 3 | 0 | 0 |

**Table S23.** Flows of grants and recorded funding from funder country to recipient country (fractional counting; data for Figure S10). [S23_country_flows]

| Funder country | Recipient country | Grants (fractional) | Domestic | USD (M, fractional) |
|---|---|---|---|---|
| United States | United States | 249 | True | 149.6 |
| United Kingdom | United Kingdom | 49.57 | True | 21.81 |
| Czechia | Czechia | 17 | True | 18.58 |
| Japan | Japan | 111 | True | 13.46 |
| Poland | Poland | 35 | True | 12.54 |
| Canada | Canada | 71 | True | 11.13 |
| Australia | Australia | 17.25 | True | 10.7 |
| Germany | Germany | 11 | True | 10.65 |
| Switzerland | Switzerland | 21.14 | True | 10.53 |
| France | France | 9.58 | True | 6.15 |
| Portugal | Portugal | 31.75 | True | 5.25 |
| European Union | France | 1.5 | False | 5.12 |
| China | China | 80 | True | 4.91 |
| Ireland | Ireland | 2 | True | 3.39 |
| European Union | Belgium | 1.7 | False | 3.17 |
| Finland | Finland | 10 | True | 3.16 |
| Denmark | Denmark | 6 | True | 2.78 |
| European Union | Netherlands | 2.33 | False | 2.45 |
| European Union | Germany | 1.25 | False | 1.7 |
| European Union | Switzerland | 2.25 | False | 1.59 |
| Israel | Israel | 2 | True | 1.43 |
| United States | Australia | 3 | False | 1.23 |
| Sweden | Sweden | 4 | True | 1.13 |
| New Zealand | New Zealand | 3 | True | 1.13 |
| European Union | United Kingdom | 1.53 | False | 1.04 |
| United Kingdom | United States | 1.08 | False | 0.9 |
| Netherlands | Netherlands | 1 | True | 0.88 |
| European Union | Ireland | 0.75 | False | 0.8 |
| United States | Taiwan | 2 | False | 0.68 |
| Slovakia | Slovakia | 5 | True | 0.66 |
| European Union | Spain | 2.67 | False | 0.66 |
| Norway | Norway | 4 | True | 0.62 |
| United Kingdom | Belgium | 0.7 | False | 0.6 |
| European Union | Denmark | 2.25 | False | 0.58 |
| India | India | 9 | True | 0.56 |
| Switzerland | United States | 0.39 | False | 0.54 |
| United Kingdom | Switzerland | 0.25 | False | 0.51 |
| United Kingdom | Germany | 1.23 | False | 0.5 |
| Switzerland | France | 0.67 | False | 0.5 |
| United Kingdom | Thailand | 0.67 | False | 0.31 |
| European Union | Portugal | 0.33 | False | 0.3 |
| United States | India | 1 | False | 0.25 |
| European Union | Greece | 0.5 | False | 0.24 |
| Australia | Canada | 0.5 | False | 0.21 |
| European Union | Austria | 1 | False | 0.2 |

| Funder country | Recipient country | Grants (fractional) | Domestic | USD (M, fractional) |
|---|---|---|---|---|
| Switzerland | Canada | 2 | False | 0.19 |
| European Union | Russia | 0.42 | False | 0.18 |
| European Union | Italy | 1 | False | 0.17 |
| European Union | South Africa | 0.67 | False | 0.17 |
| European Union | Israel | 1 | False | 0.17 |
| Spain | Spain | 2 | True | 0.17 |
| Switzerland | Georgia | 1 | False | 0.16 |
| United Kingdom | Brazil | 0.33 | False | 0.15 |
| United Kingdom | Uganda | 0.5 | False | 0.15 |
| European Union | United States | 0.5 | False | 0.14 |
| European Union | Georgia | 1.15 | False | 0.13 |
| United Kingdom | Italy | 0.2 | False | 0.13 |
| United Kingdom | Australia | 0.2 | False | 0.12 |
| Switzerland | Belgium | 0.4 | False | 0.12 |
| Switzerland | Germany | 0.4 | False | 0.12 |
| Switzerland | Singapore | 1 | False | 0.11 |
| United Kingdom | Denmark | 0.2 | False | 0.11 |
| United Kingdom | France | 0.2 | False | 0.11 |
| Portugal | France | 1.12 | False | 0.11 |
| Australia | United States | 0.25 | False | 0.1 |
| United States | United Kingdom | 1 | False | 0.1 |
| United Kingdom | Canada | 0.5 | False | 0.09 |
| United Kingdom | Tanzania | 0.25 | False | 0.07 |
| France | Belgium | 0.58 | False | 0.05 |
| United Kingdom | Hungary | 0.12 | False | 0.05 |
| China | Canada | 0.5 | False | 0.05 |
| Portugal | Netherlands | 0.5 | False | 0.04 |
| Portugal | Spain | 0.5 | False | 0.04 |
| European Union | Ukraine | 0.2 | False | 0.04 |
| France | Denmark | 0.25 | False | 0.03 |
| France | United States | 0.25 | False | 0.03 |
| China | Singapore | 1 | False | 0.03 |
| France | Norway | 0.33 | False | 0.02 |
| Latvia | Latvia | 1 | True | 0.02 |
| Portugal | Sweden | 0.12 | False | 0.02 |
| China | United States | 0.5 | False | 0.02 |
| United Kingdom | Ireland | 1 | False | 0 |

**Table S24.** Concentration of recorded funding: Herfindahl–Hirschman index (0–10,000), Gini coefficient and top 10% share, for funders, funder countries, recipient countries and recipient institutions. [S24_concentration]

| Unit | N | HHI (0-10,000) | Gini | Top 10% share (%) | Largest unit | Largest share (%) |
|---|---|---|---|---|---|---|
| Funders | 105 | 610 | 0.696 | 51.7 | National Institute of Allergy and Infectious Diseases | 21 |
| Funder countries | 25 | 2,501 | 0.707 | 55.7 | United States | 47.7 |
| Recipient countries (fractional) | 39 | 2,461 | 0.789 | 64.7 | United States | 47.5 |
| Recipient institutions (fractional) | 525 | 78 | 0.683 | 53.4 | Intralytix (United States) | 2.7 |

**Table S25.** Shares of recorded funding from US funders and from NIAID, and funder HHI, by period. [S25_shares_by_period]

| Period | Grants | Total (USD M) | US funders (%) | NIAID (%) | Funder HHI |
|---|---|---|---|---|---|
| 1999-2008 | 102 | 29.1 | 58.1 | 13.6 | 726 |
| 2009-2015 | 198 | 76.6 | 44.1 | 17.4 | 674 |
| 2016-2023 | 502 | 213 | 47.5 | 23.3 | 718 |
| All years | 802 | 318.6 | 47.7 | 21 | 610 |

**Table S26.** Collaboration counts: grants with several recipient institutions or countries, and grants linking Global North and Global South recipients.

[S26_collaboration]

| Measure | Count | Share of grants (%) |
|---|---|---|
| Grants with more than one recipient institution | 135 | 16.8 |
| Grants with recipients in more than one country | 39 | 4.9 |
| Distinct country pairs in those grants | 61 | |
| Grants linking Global North and Global South recipients | 7 | 0.9 |
| Grants linking high-income and low or lower-middle income recipients | 2 | 0.2 |
| Grants with 2 or more named researchers | 186 | 23.2 |
| Co-investigator pairs | 2045 | |
| Pairs that worked together on 2 or more grants | 73 | |

**Table S27.** Growth, US share and concentration with and without NIAID grants. [S27_without_niaid]

| Sample | Grants | Total (USD M) | Grant growth (%/yr) | Grant growth CI | Funding growth (%/yr) | Funding growth CI | US funders (%) | Funder HHI | Funder-country HHI |
|---|---|---|---|---|---|---|---|---|---|
| All funders | 802 | 318.6 | 12.2 | 10.0 to 14.5 | 16.6 | 12.6 to 20.7 | 47.7 | 610 | 2,501 |
| Without National Institute of Allergy and Infectious Diseases | 721 | 251.8 | 11.7 | 9.5 to 14.1 | 16.2 | 11.7 to 20.9 | 33.8 | 272 | 1,508 |

**Table S28.** Annual growth of recorded funding in nominal and constant 2023 US dollars (CPI-U). [S28_real_growth]

| | Annual growth (%) | 95% CI low | 95% CI high | Years | Total (USD M) |
|---|---|---|---|---|---|
| Nominal USD | 16.6 | 12.6 | 20.7 | 1999-2022 | 318.6 |
| Constant 2023 USD (CPI-U) | 14.1 | 10.2 | 18.2 | 1999-2022 | 396.1 |

**Table S29.** Screening flow from the Dimensions export to the analysed grants (data for Figure 6 / Figure S11). [S29_screening_flow]

| Step | Records | Note |
|---|---|---|
| Records exported from Dimensions (13 Feb 2024) | 1521 | |
| Excluded: not about therapeutic use of phages | 391 | 386 coded 0, 5 coded -1 |
| Relevant grants | 1130 | |
| Excluded from dollar analyses: no amount in Dimensions | 312 | kept for count-based checks |
| Grants with an amount | 818 | |
| Excluded: start year outside 1999-2023 | 16 | |
| Analysed (dollar analyses) | 802 | |
| Relevant grants without an amount, 1999-2023 (count-based checks) | 272 | |

**Table S30.** Award types among relevant grants with and without a recorded amount. [S30_award_types]

| Award type | With an amount (n) | Without an amount (n) | With an amount (%) | Without an amount (%) |
|---|---|---|---|---|
| Research grant | 717 | 136 | 89.4 | 50 |
| Training award | 61 | 117 | 7.6 | 43 |
| Capacity funding | 24 | 19 | 3 | 7 |
| Total | 802 | 272 | 100 | 100 |

**Table S31.** Count-based shares (% of grants) with and without the 272 grants that have no amount. [S31_count_based_shares]

| Measure (share of grants) | With an amount | Without an amount | All relevant grants | All, without training awards |
|---|---|---|---|---|
| Grants | 802 | 272 | 1,074 | 896 |
| Recipients in the Global South (%) | 11.8 | 34.4 | 17.5 | 14.1 |
| Recipients in low or lower-middle income countries (%) | 1.4 | 1.7 | 1.4 | 1.7 |
| Recipients with no country (%) | 0 | 17.3 | 4.4 | 3.8 |
| Recipients in the United States (%) | 31.4 | 11.2 | 26.3 | 29.6 |
| Recipients in Brazil (%) | 0 | 28.7 | 7.3 | 2.4 |
| Funders in the Global South (%) | 11.3 | 39 | 18.3 | 14.3 |
| Funders in the United States (%) | 31.9 | 12.1 | 26.9 | 30.2 |
| Growth in grants per year, 1999-2022 (%) | 12.2 | | 13.1 | 13 |

**Table S32.** Shares of funding with imputed amounts for grants without an amount (low, central and high scenarios). [S32_imputed_amount_scenarios]

| Measure | Recorded amounts only (main analysis) | Imputed, low (25th percentile) | Imputed, central (median) | Imputed, high (75th percentile) |
|---|---|---|---|---|
| Grants with an amount | 802 | 1,074 | 1,074 | 1,074 |
| Total (USD M) | 318.6 | 331.8 | 353.8 | 400.1 |
| Recipients in the Global South (%) | 2.11 | 2.64 | 3.8 | 5.49 |
| Recipients in low or lower-middle income countries (%) | 0.33 | 0.55 | 0.54 | 0.52 |
| Recipients in Brazil (%) | 0.05 | 0.32 | 1.23 | 2.64 |
| Funding from US funders (%) | 47.65 | 47.4 | 45.11 | 41.39 |
| Funding from NIAID (%) | 20.98 | 20.15 | 18.9 | 16.71 |
| Funder-country HHI | 2,501 | 2,466 | 2,245 | 1,953 |
| Funding growth 1999-2022 (%/yr) | 16.6 | 16.3 | 16 | 16.1 |

**Table S33.** Yearly grant records merged into projects (same funder, same translated title, consecutive years). [S33_projects]

| Measure | Grant records | Projects |
|---|---|---|
| Units | 802 | 771 |
| Units built from more than one record | 0 | 25 |
| Growth in units per year, 1999-2022 (%) | 12.2 | 12.3 |
| Transient researchers, start-year rule (%) | 82.1 | 82.9 |
| Transient researchers, full-span rule (%) | 3.5 | 3.5 |

**Table S34.** Female-only vs male-only grant size gap in subsets of grants. [S34_gender_subsets]

| Subset | Grants in model | Female-only vs male-only (%) | 95% CI low (%) | 95% CI high (%) | p |
|---|---|---|---|---|---|
| All grants (main result) | 742 | -18.731 | -32.289 | -2.458 | 0.026 |
| Without funders in Japan and China | 549 | -20.864 | -36.58 | -1.253 | 0.038 |
| Without training awards | 686 | -20.224 | -34.176 | -3.315 | 0.021 |

**Table S35.** Topic shares of funding from the stored assignment, from the refitted model, and split by topic probability. [S35_topic_probability_shares]

| Topic | One topic per grant, paper assignment (%) | One topic per grant, refit (%) | Split by topic probability, refit (%) | 1999-2005, paper assignment (%) | 1999-2005, split, refit (%) | 2016-2023, paper assignment (%) | 2016-2023, split, refit (%) |
|---|---|---|---|---|---|---|---|
| Infectious disease control | 35.9 | 32.7 | 30.6 | 23 | 23.3 | 39.4 | 32.9 |
| Bacterial genetics and mechanisms | 28.8 | 14.7 | 15.9 | 23.8 | 31 | 25.2 | 12.3 |
| Antibiotic resistance strategies | 22.4 | 41.2 | 39.7 | 2.8 | 11.8 | 25.7 | 44.3 |
| Immunotherapy and targeted treatments | 12.9 | 11.3 | 12.7 | 50.4 | 33.4 | 9.6 | 9.4 |
| Unclassified | 0 | 0.1 | 1.1 | | 0.5 | 0 | 1.1 |

**Table S36.** Headline results under each sensitivity check (data for Figure S12). [S36_robustness_summary]

| Check | Funding growth (%/yr) | Global South recipients (% of funding) | Low + lower-middle income recipients (%) | US funders (% of funding) | NIAID (% of funding) | Female-only grant gap (%) | One-year-only researchers (%) | Rise in antibiotic-resistance share, 1999-2005 to 2016-2023 (points) |
|---|---|---|---|---|---|---|---|---|
| Main analysis | 16.6 | 2.1 | 0.33 | 47.7 | 21.0 | -18.73 | 82.1 | 22.9 |
| Missing amounts: imputed, low (25th percentile) | 16.3 | 2.64 | 0.55 | 47.4 | 20.15 | | | |
| Missing amounts: imputed, central (median) | 16.0 | 3.8 | 0.54 | 45.11 | 18.9 | | | |
| Missing amounts: imputed, high (75th percentile) | 16.1 | 5.49 | 0.52 | 41.39 | 16.71 | | | |
| Count of grants instead of dollars (all 1,074) | | 17.5 | 1.4 | | | | | |
| Projects instead of records | | | | | | | 82.9 | |
| Full-span engagement rule | | | | | | | 3.5 | |

| Check | Funding growth (%/yr) | Global South recipients (% of funding) | Low + lower-middle income recipients (%) | US funders (% of funding) | NIAID (% of funding) | Female-only grant gap (%) | One-year-only researchers (%) | Rise in antibiotic-resistance share, 1999-2005 to 2016-2023 (points) |
|---|---|---|---|---|---|---|---|---|
| Researchers first seen 2001-2019 | | | | | | | 83.31 | |
| Researchers who held only one grant | | | | | | | 83.9 | |
| Amounts spread over grant years | 17.1 | | | | | | | |
| Income group at grant start | | | 0.46 | | | | | |
| OECD DAC list instead of Global South | | 2.17 | | | | | | |
| Constant 2023 dollars | 14.1 | | | | | | | |
| Without NIAID | 16.2 | | | 33.8 | | | | |
| Gender: without funders in Japan and China | | | | | | -20.86 | | |
| Gender: without training awards | | | | | | -20.22 | | |
| Gender: amount per year | | | | | | -21.65 | | |
| Gender: simple model | | | | | | -27.28 | | |
| Topics: refit model, split by probability | | | | | | | | 32.5 |
| Conclusion holds if | > 10 | < 10 | < 1 | > 30, largest country | > 15, largest funder | < 0 | > 50 | > 10 |

**Table S37.** Grants and recorded funding by research topic. [Table3_topics]

| Topic | Grants | Total (USD M) | Median (USD) | Share of funding (%) |
|---|---|---|---|---|
| Infectious disease control | 280 | 114.5 | 194,706 | 35.9 |
| Bacterial genetics and mechanisms | 237 | 91.7 | 171,755 | 28.8 |
| Antibiotic resistance strategies | 151 | 71.4 | 242,263 | 22.4 |
| Immunotherapy and targeted treatments | 133 | 41 | 111,442 | 12.9 |
| Unclassified | 1 | 0 | 49,400 | 0 |

**Table S38.** Share of recorded funding by World Bank income group (fiscal year 2024), for funders and recipients (fractional counting). [Table5_income_groups]

| | Funders (%) | Recipients, fractional (%) |
|---|---|---|
| High income | 98.25 | 97.78 |
| Upper middle income | 1.57 | 1.89 |
| Lower middle income | 0.18 | 0.29 |
| Low income | 0 | 0.05 |
